# Development of a resilience assessment tool for cardiac care pathways in Europe: A mixed-methods study

**DOI:** 10.1101/2025.09.01.25334825

**Authors:** Ana Sofia V. Carvalho, Óscar Brito Fernandes, Jan J. Piek, Josepa Mauri, Ariadna Sanz Escartín, William Wijns, Niek Klazinga, Dionne Kringos

## Abstract

**Objectives:** To develop a resilience assessment tool for cardiac care pathways, informed by stakeholder insights on the impact of COVID-19, emerging innovations, and recommendations to enhance resilience.

**Design:** Mixed-methods study comprising three phases: 1) survey among European cardiac care providers; 2) five multi-stakeholder focus groups; 3) collaborative tool development. Quantitative data were analysed using descriptive statistics, and qualitative data were analysed thematically.

**Setting:** The survey targeted cardiology professionals from the 27 EU Member States and Ukraine who worked during the COVID-19 pandemic. Focus group participants were purposefully sampled to represent clinical, organisational, and policy perspectives.

**Participants:** A total of 177 survey respondents and 40 informants in focus groups.

**Results:** Six key resilience dimensions of cardiac care pathways were identified: workforce, organisation of care delivery, governance and trust, communication and cooperation, medical devices and products, and data collection and use. Staff shortages and infrastructure capacity were key challenges during the pandemic. The most frequent measures were the reallocation of health staff (75%; n=133) and repurposing infrastructures (38%; n=32). Participants discussed the six resilience dimensions around a total of 17 sub-dimensions and 39 recommended actions to enhance resilience were identified. The resulting resilience assessment tool included four components: 1) mapping a context-specific cardiac care pathway; 2) stakeholder identification critical to participate in collective self-assessment; 3) a preparedness checklist generating a visual heat map; and 4) a resource toolkit.

**Conclusions:** The resilience assessment tool offers step-by-step guidance to strengthening cardiac care pathways across six key resilience dimensions, supported by actionable recommendations. The tool enables the identification of context-specific vulnerabilities and improvement priorities, thereby supporting healthcare professionals and policymakers in enhancing preparedness and ensuring care continuity before, during, and after crises. Its implementation is currently being piloted in European hospitals to evaluate and refine its practical applicability.

**ARTICLE SUMMARY:** **Strengths and limitations of this study**

- This study draws on the lessons from COVID-19, informed by stakeholder insights, to fill the gap of systematic tools tailored to enhance the resilience of cardiac care pathways.
- A sequential mixed-methods approach aimed to foster cross-country learning and the identification of key challenges and strategies to enhance cardiac care resilience.
- Despite a low survey response rate, the combination of survey data with insights from focus groups and a co-creation process, supported the development of a context-specific resilience assessment tool for cardiac care pathways.
- The continuous involvement from the RESIL-Card Consortium and Advisory Board ensured the tool’s relevance and usability, to support policymakers and healthcare professionals in identifying and prioritising context-specific vulnerabilities in cardiac care.
- While the tool is specifically tailored to cardiac care, future research could test and adapt these findings to other clinical areas and geographies.

## 1. BACKGROUND

The COVID-19 pandemic caused major disruptions in cardiac healthcare delivery worldwide, notably reductions in the number of diagnoses of acute and chronic conditions, delays in seeking care and in access to treatment [1, 2, 3], resulting in higher mortality rates from heart diseases [3, 4, 5]. These consequences add to the large burden that cardiovascular diseases pose to health systems across Europe, as the leading cause of mortality [6] and a relevant cause of disability [7]. Furthermore, the pandemic had a significant impact on healthcare workers’ mental health and aggravated workforce shortages [8].

The impact of the COVID-19 pandemic on cardiac care delivery and outcomes underscores the pressing need to improve care pathways’ resilience to future systemic shocks, such as extreme climatic events, war conditions, cyberattacks, or financial crises, considering the constantly evolving political, economic, and climatic context worldwide. Health systems inputs generally include components such as the health workforce, physical infrastructure, and financial resources [9], while service delivery can be defined as ‘the combination of inputs into a production process that takes place in a particular organizational setting and that leads to the delivery of a series of interventions’ [10]. While the COVID-19 crisis has triggered swift adaptations and innovations in care delivery, there is a growing recognition that resilience should be embedded in systematic health system performance assessment and not only in crisis response [11, 12, 13].

Resilience can be defined as the ‘institutions’ and health actors’ capacities to prepare for, recover from and absorb shocks, while maintaining core functions and serving the ongoing and acute care needs of their communities’ [14]. International reports have provided recommendations on how to improve health systems resilience [15, 16], notably focusing on establishing frameworks for coordination and decision-making prior to crises and anticipating the required capabilities and resources [15]. Nonetheless, there is a lack of systematic tools tailored to the cardiac care pathway, particularly taking stock of the lessons learned from the COVID-19 crisis.

The RESIL-Card Consortium was formed to address this critical gap (https://www.wecareabouthearts.org/resil-card/the-project/), consisting of four partners: We CARE Alliance (France), the Amsterdam UMC Health Care Systems and Services Research Group (Netherlands); the Catalan Health Service - CatSalut (Spain); and the Italian Society of Interventional Cardiology (GISE, Italy). It draws on the experiences of health system actors in the cardiology ecosystem, across clinical, organisational and policy levels, to learn how to strengthen cardiac care pathways’ resilience and to foster cross-learning within and across organisations and countries.

This study aimed at developing a resilience assessment tool for cardiac care pathways. The tool intends to provide a structured and adaptable approach for stakeholders in the cardiology ecosystem to assess their preparedness and enhance resilience within their contexts. The development of the tool was guided by three research questions, from the perspective of stakeholders in the cardiology ecosystem:

1. How did the COVID-19 pandemic disrupt cardiac care inputs and care delivery process?
2. What organisational practices and innovations emerged during the COVID-19 pandemic, and to what extent did they contribute to the continuity and quality of cardiac care?
3. What actions do stakeholders recommend to strengthen the resilience of cardiac care pathways for future health system shocks?

## 2. METHODS

### Study design and setting

We conducted an explanatory sequential mixed-methods study [17] comprising three phases: a survey followed by multi-stakeholder focus groups, which culminated in the development of a resilience assessment tool (Figure 1). This approach was conducted to obtain a comprehensive understanding of the COVID-19 pandemic experiences from a diversity of stakeholders and to develop a tool based on these experiences. This study adheres to the requirements of mixed-methods studies by Lee et al [18] (See Supplementary File 1).

**Figure 1.**
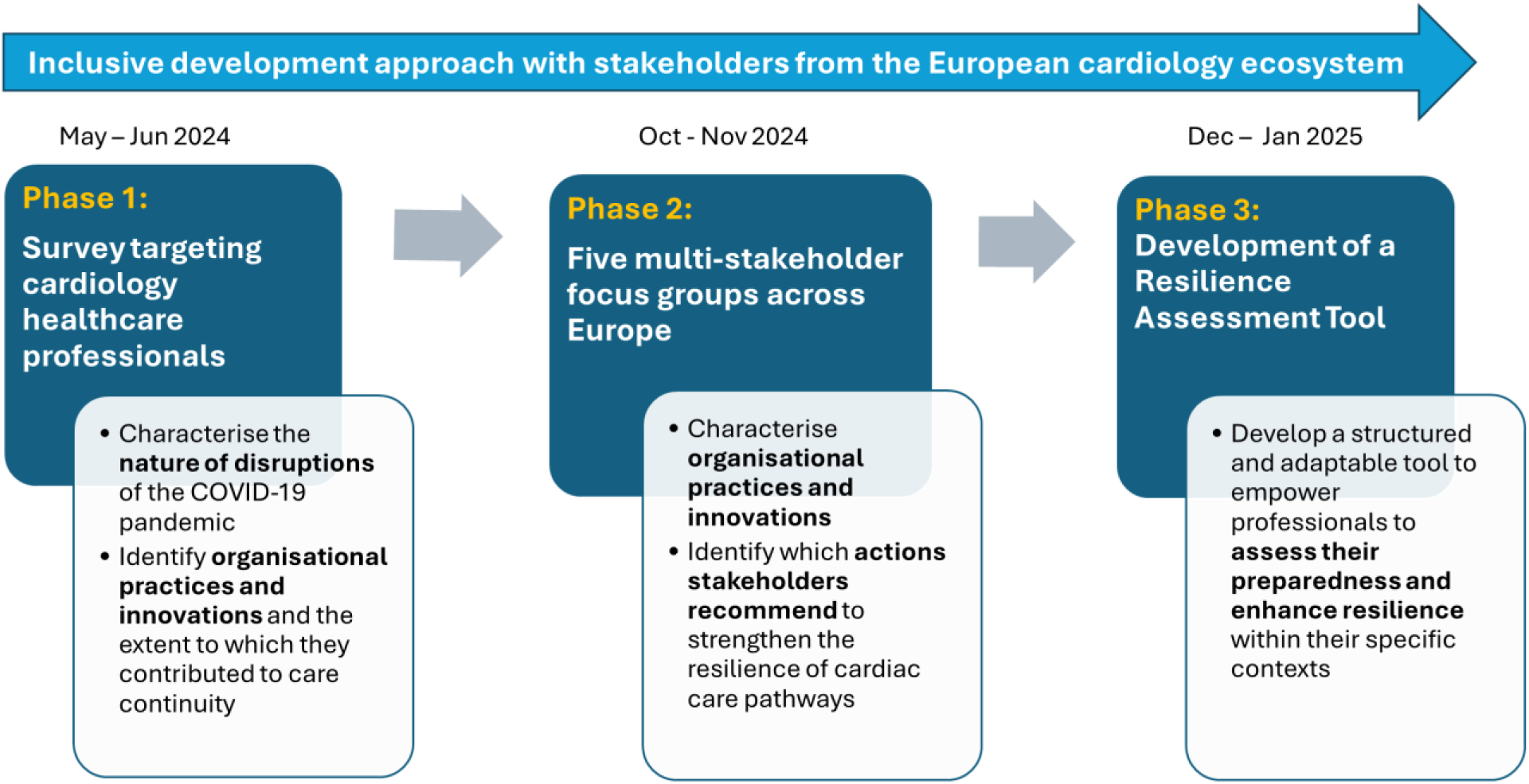
Overview of the research phases to develop a resilience assessment tool for cardiac care pathways.

First, a survey (Phase 1) was distributed to European cardiology healthcare professionals, aiming to identify and characterise the nature of the cardiac care disruptions caused by the COVID-19 pandemic, organisational practices and innovations. Second, informed by the survey’s results, five focus groups (Phase 2) involving cardiac care stakeholders across European countries were conducted, to discuss the survey’s results and complement where needed. Quantitative data guided the qualitative data collection and was later integrated during the data analysis. The qualitative findings helped to interpret the quantitative results and provided additional insights. Third, informed by Phases 1 and 2, a Resilience Assessment Tool for cardiac care pathways (Phase 3) was developed involving a broader research team – the RESIL-Card Consortium –, supported by a multi-stakeholder Advisory Board, including patient representatives.

### Data collection

#### Phase 1: Survey targeting cardiac care providers

A survey was developed, targeting healthcare professionals from the cardiology field from the 27 EU Member States and Ukraine who had worked during the COVID-19 pandemic. It was informed by a scoping review published by some of the authors on the impact of the COVID-19 pandemic on cardiac care pathways [2] and on work from the We CARE Alliance on the impact and adaptations in care delivery during the pandemic [19] (Figure 2). In addition, relevant scientific literature and international reports on health systems resilience were consulted [15] [20]. The first iteration of the survey received input from four cardiologists from the RESIL-Card Consortium and from Advisory Board members. Following adjustments to cardiology-related terms and additional questions on gender, the questions were discussed in four face-to-face cognitive testing interviews (Supplementary File 2). The survey was piloted on the intended online platform (Microsoft Forms) with 11 people from the RESIL-Card Consortium, providing an estimation of the average time required to complete the survey (24 minutes).

**Figure 2.**
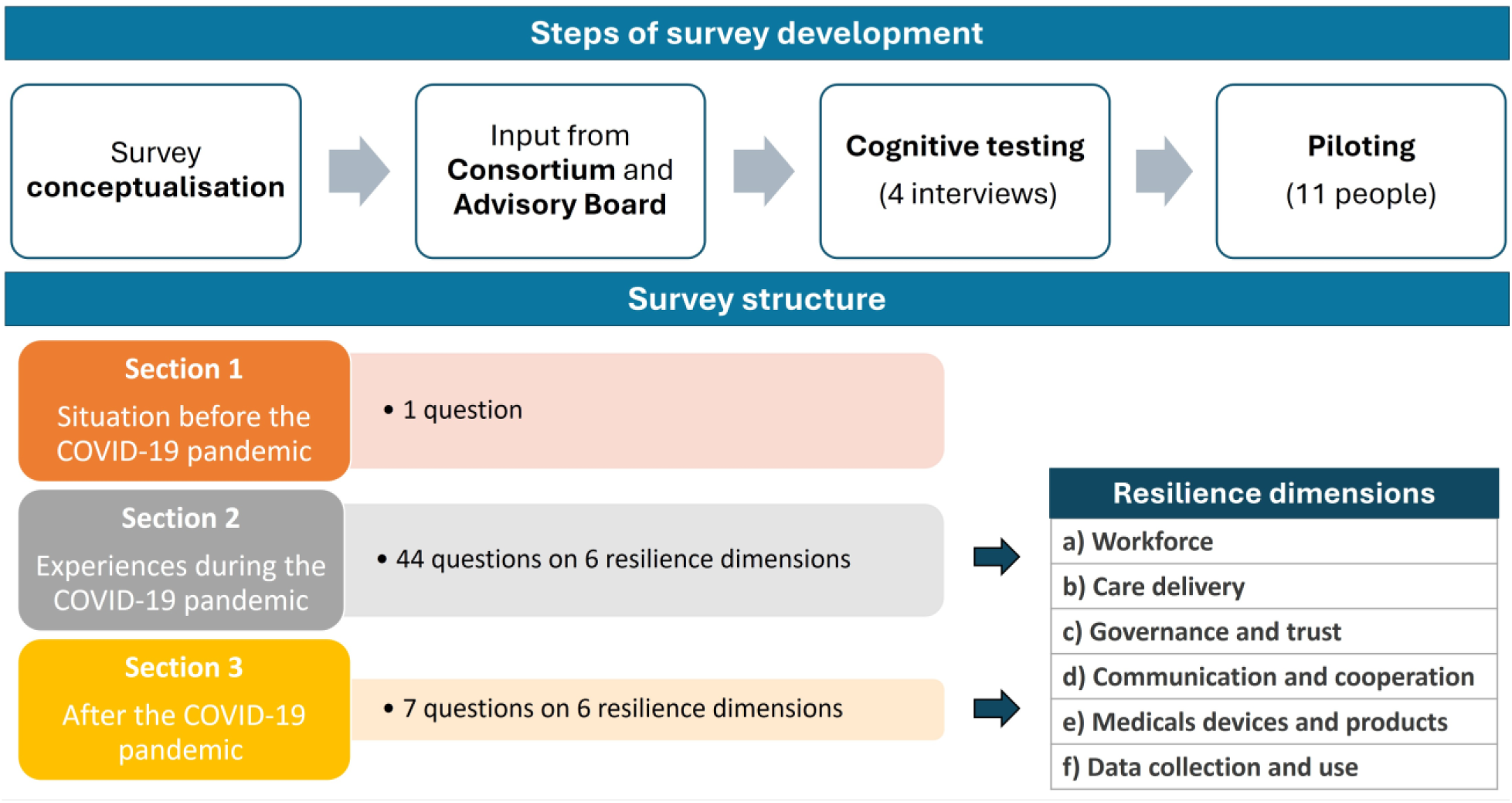
Steps of the survey development and survey’s structure.

The survey was structured in three sections: 1) situation before the COVID-19 pandemic, 2) experiences during the pandemic, and 3) lessons learned after the pandemic. The first section included one question to rate the most relevant challenges before the pandemic. The sections two and three were organised in six sub-sections, which were identified from the literature as relevant dimensions to strengthen resilience: (a) workforce, (b) organisation of care delivery to cope with surge in demand, (c) governance and trust, (d) communication and cooperation, (e) medical devices and products, and (f) data collection and use. Supplementary File 3 presents the final version of the survey, which included 52 open- and closed-ended questions.

The data collection comprised a period of six weeks, between 5^th^ May and 16^th^ June 2024. A link to the questionnaire was sent via email to 4338 European members of ‘PCR companions’ community, an international network of interventional cardiologists and allied professionals, followed by one reminder and reference to the survey in two additional communications. Various partner organisations contributed to the dissemination of the questionnaire, including Societies of Cardiology from the targeted countries. Several initiatives were developed to raise awareness of the survey in national congresses and in social media (Supplementary File 4).

#### Phase 2: Multi-stakeholder focus groups

A total of five focus groups were organised, comprising between four and eleven participants. A purposeful sample of stakeholders was selected for the focus groups, representing three levels of health systems decision-making: micro (clinical), meso (organisational), and macro (policy). First, four focus groups convened, with participants from the countries most represented in the survey responses: two Italian focus groups with a regional focus - Campania and Lombardy regions - and two focus groups with a national focus – Spain and Netherlands. To discuss the transferability of these findings to other European countries, an international focus group was organised, with participants from Belgium, Greece, Poland, Portugal, Romania, and Sweden.

Each RESIL-Card Consortium partner was responsible for selecting and inviting participants by email in their native language (Supplementary File 5). The invitations for the international focus group aimed for a balanced geographical distribution across EU countries with participants proficient in English. Pre-reading materials in English were sent to all focus groups participants, including information about the project, the discussion questions, and presentation of the moderators.

The focus groups were conducted online and lasted on average 110 minutes. National and regional focus groups were held in the participants’ native languages — Italian, Spanish, and Dutch — and moderated by each Consortium partner in their respective native languages. This allowed participants to express themselves without a language barrier. The international focus group was conducted in English. The meetings were recorded and transcribed verbatim with the participants’ consent.

The focus group was structured in three parts. First, ASVC (first author) presented the project’s aims and key findings from the survey. Second, the moderator guided the discussion around each of the six resilience dimensions addressed in the survey, focusing on two questions: 1) During the COVID-19 pandemic, what was your experience with practices and innovations implemented to ensure continuity of cardiac care? 2) What could be done differently in a future crisis to be more prepared and effective? Third, the conceptualisation of the resilience assessment tool was discussed, focusing on two aspects: users and format. Finally, participants were asked about their interest to be involved in the piloting phase of the resilience tool.

#### Phase 3: Co-development of the resilience assessment tool

To conceptualise the tool, the authors considered the insights from the focus groups participants, as well as examples of resilience tools from the literature (such as [21, 22]). An inclusive, multi-stakeholder approach was conducted, with ongoing contributions of the RESIL-Card Consortium and Advisory Board during various moments of consultation.

### Data analysis

#### Survey data

Quantitative data were analysed using descriptive statistics, conducted with Microsoft Excel. These data informed our research questions, namely: 1) the nature of the disruptions during the COVID-19 pandemic, 2) practices implemented, and 3) the prioritisation of actions to strengthen the resilience of cardiac care pathways. The qualitative data from the survey’s open-ended questions was analysed using thematic analysis [23], through inductive coding. These data informed the third research question, namely by identifying themes to strengthen resilience, which were considered as preliminary themes for discussion in the focus groups.

#### Focus groups data

The raw data comprised video recordings and transcriptions of focus group discussions with experts, and meeting notes prepared by each focus group moderator. The moderators of the focus groups reviewed the transcripts in their native language and translated them into English. A thematic analysis approach following the Template Analysis procedure [24] was used to analyse the data [25], using an inductive approach, considering a priori themes identified in the survey.

A stepwise process was conducted, as follows: 1) familiarisation with the data, conducting preliminary coding, supported by the summaries of the focus groups and the themes derived from the survey across the six resilience dimensions; 2) development of a code book to systematically categorise the data (appendix); 3) coding of focus groups transcripts according to the code book. The coding process was conducted by ASVC, adhering to the code book, and reviewed by the other authors. The code book was continuously refined throughout the coding process. In step 4, themes and sub-themes were identified from the data.

The preliminary results from the first three focus groups (Spain and Italy) were discussed with the Consortium during the project’s General Assembly in October 2024. These discussions helped to refine themes and informed the topics to be explored in the subsequent focus groups. After the coding of all focus groups transcripts, team discussions were convened to refine the themes and sub-themes, including three one-hour meetings between ASVC, OBF, NK, and DK as well as a one-hour Consortium meeting.

#### Synthesis of data to develop the resilience assessment tool

The themes identified in the survey and focus groups were complemented and refined to develop the tool: each theme was considered a ‘sub-dimension of resilience’, representing key aspects to be addressed within each dimension. For each resilience sub-dimension, sub-themes were proposed as specific actions to be undertaken by stakeholders. Based on the key actions identified, questions for the preparedness checklist and specific recommendations were developed. The checklist questions and recommendations were reviewed and refined in discussions within the AUMC research team and the Consortium.

#### Patient and public involvement

Patient representatives participated in all RESIL-Card Consortium meetings, being closely involved in all research phases of this study. As members of the RESIL-Card Advisory Board, patient representatives participated in the survey conceptualisation and dissemination. Patients and/or patient representatives were involved as informants in four of the five focus groups conducted. The tool was reviewed by patient representatives.

## 3. RESULTS

### Study population

A total of 177 healthcare professionals responded to the survey (Supplementary File 6). Most of the respondents were physicians (92%, n=163), specifically interventional cardiologists (70% of the respondents; n=124), followed by general cardiologists (15%, n=27). Seven percent of the respondents were nurses (n=13). Professionals worked mostly in Italy (27%, n=47) and Spain (24%, n=43), in public hospitals (86%, n=153) and in hospitals with cardiac catheterisation laboratory available on a 24/7 basis (47%, n=83).

A total of 40 informants participated in the five focus groups, with a range between 4 and 11 participants per focus group, from nine European countries. Informants represented the three health system levels and a broad range of stakeholder type (Table 1) (see Supplementary File 7 for a detailed breakdown of participants).

**Table 1.**
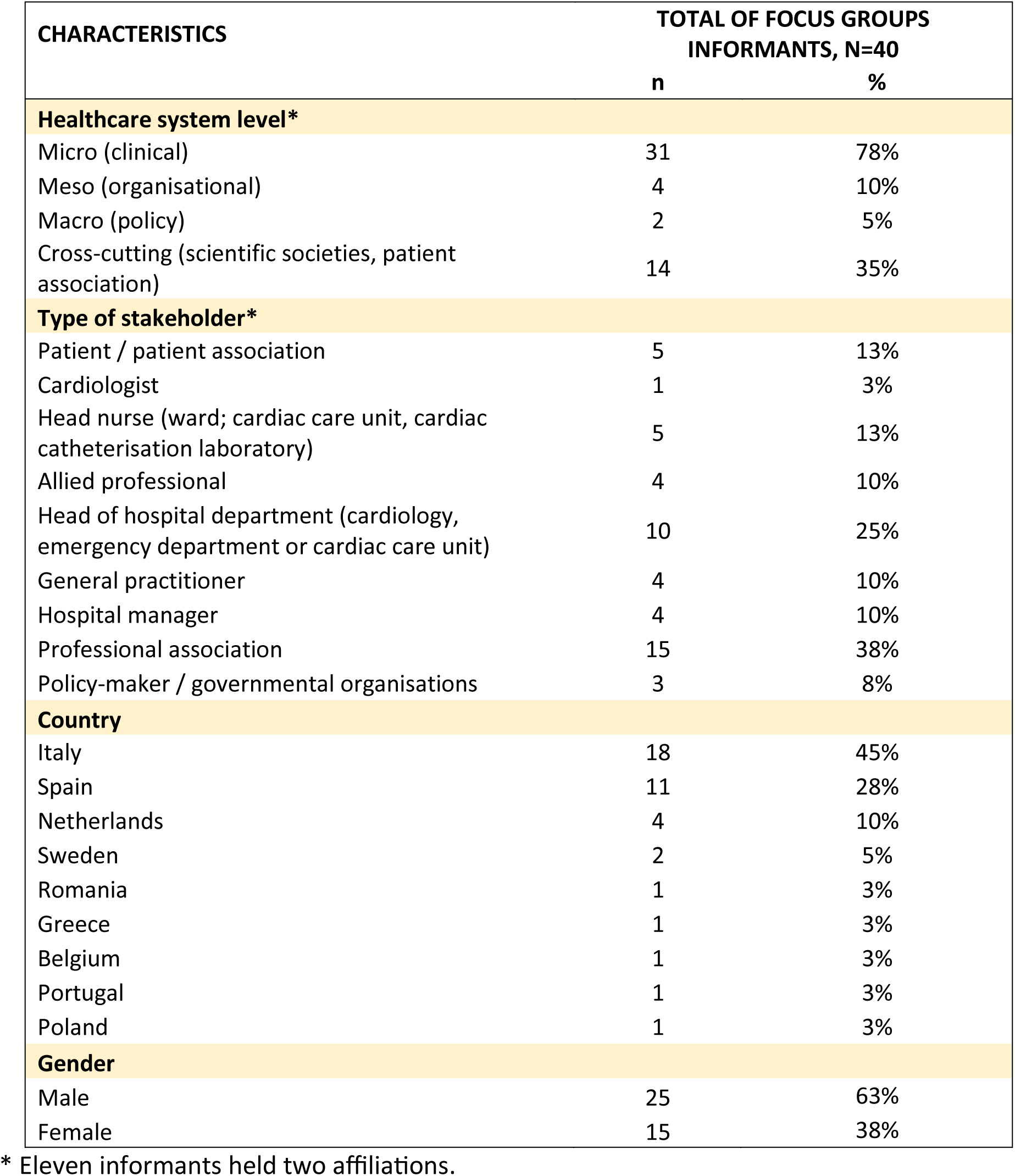
Summary of the characteristics of the focus groups’ informants (n=40)

### Nature of disruptions to service delivery inputs and care delivery processes, from the perspective of European cardiac care providers

Infrastructure capacity and staff shortages were perceived as the most relevant challenges to provide cardiac care during the COVID-19 pandemic: shortage of infrastructure capacity was perceived as very important or extremely important by 38% of respondents (n=68) and staff shortages due to SARS-CoV-2 infections by 35% (n=62) (Figure 3 (a)). Infrastructures and workforce shortages were also perceived as the most relevant challenges before the pandemic (see Supplementary File 8 for a detailed breakdown).

**Figure 3.**
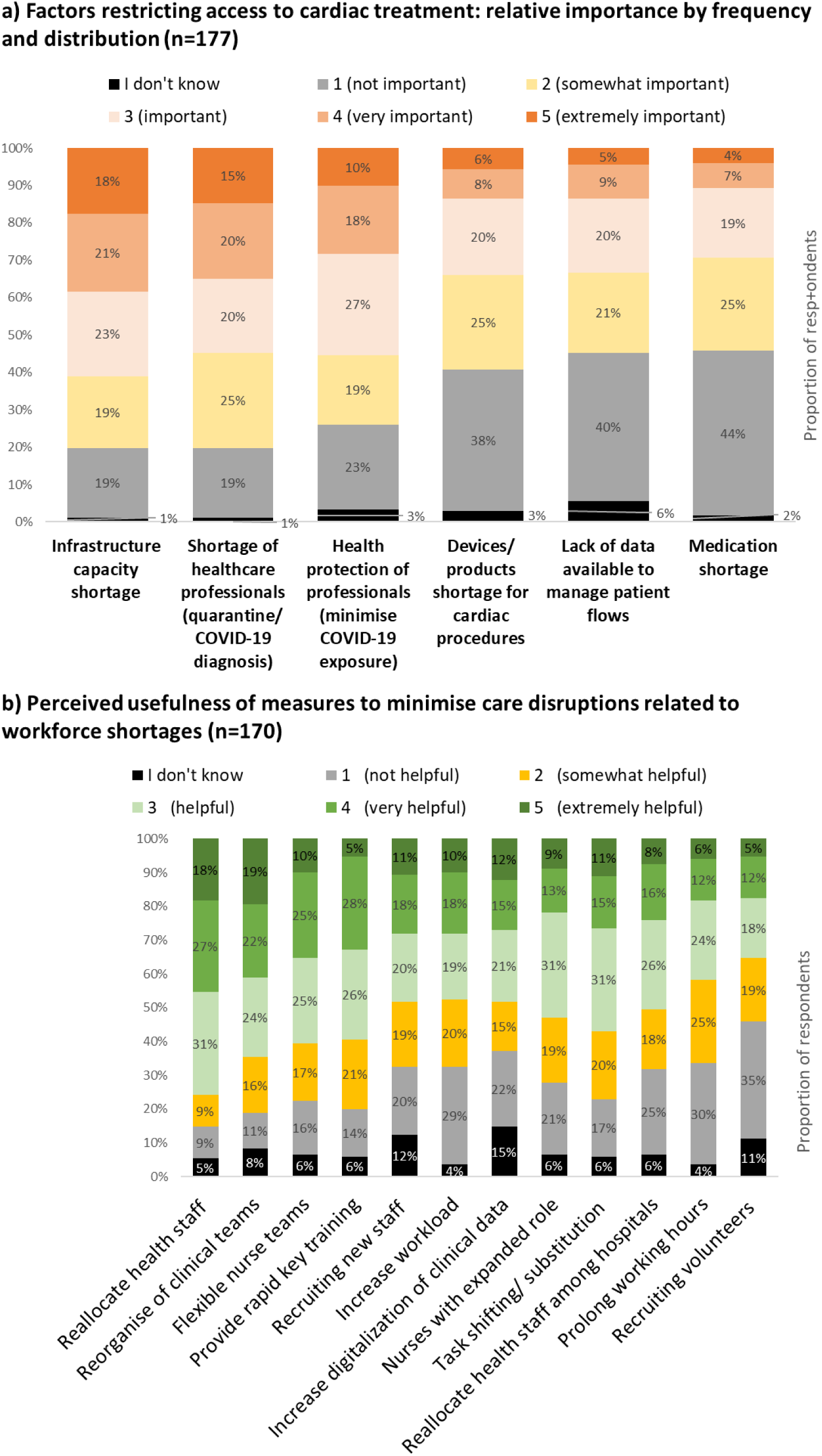
Factors restricting access to cardiac treatment and the perceived usefulness of measures to minimise acute cardiac care disruptions caused by workforce shortages.

During the COVID-19 pandemic, the most frequent shortage in infrastructure capacity reported was related to ICU beds (n=152, 73%) (see Supplementary File 9 for a detailed analysis). Delays in accessing diagnostic procedures were frequently reported (n=116, 66%). The therapeutic procedure with waiting times most frequently reported, from a clinical perspective, was elective diagnostic cardiac catheterization (n=120, 68%). Regarding communication and cooperation aspects, changes in referral policies were frequently mentioned (n=95; 54%). The professionals most frequently involved and responsible for decision-making were hospital managers and the heart team, with varying percentages depending on the decisions. Concerning data aspects, 45% of the respondents (n=81) reported that data was available to monitor the volume of patients with cardiac diseases by each hospital. Shortages in essential medical devices were reported by 49% of the respondents (n=86), being more frequently related to mechanical ventilators (n=77; 47%).

### Practices and innovations during the COVID-19 pandemic and its perceived usefulness

The measure most frequently reported to address staff shortages during the COVID-19 pandemic was the reallocation of health staff to key clinical areas, mentioned by 75% of respondents (n=133) (Supplementary File 9 presents the complete list and frequency of measures implemented). Furthermore, reallocation of health staff was considered the most useful practice to minimise cardiac care disruptions, reported as very or extremely helpful by 45% of respondents (n=77), followed by the reorganisation of clinical teams to spread expertise (41%, n=70), the availability of flexible nurse teams (35%, n=60) and providing rapid training in key clinical areas (33%, n=56) (Figure 3 (b)).

To address infrastructure capacity shortages at the hospital level, the most frequent practices were repurposing infrastructures (38%, n=32) and reducing regular activity (26%, n=22). New care delivery models were reported by 37% of the respondents (n=66), while 54% respondents (n=96) reported that no innovation in care delivery models was observed. Public awareness campaigns to citizens were reported by 57% of respondents (n=101) and new communication channels to inform patients were implemented 43% of the cases (n=76). A total of 55% of the respondents (n=98) reported decision-making processes through meetings, with inclusion of physicians. We observed no difference in trends between Italian and Spanish respondents, regarding the most frequent practices implemented during the COVID-19 pandemic.

### Actions recommended to strengthen the resilience of cardiac care pathways

Workforce’s health protection and enhanced communication with patients were considered as very or extremely important factors for ensuring access to cardiac care in future crises by 69% of survey respondents (n=122). The proportion of respondents who considered other factors as very or extremely important, across the six resilience dimensions, ranged between 62% (n=110) concerning improving coordination with primary care providers and 41% (n=73) in relation to improving international cooperation. Supplementary File 10 provides a detailed breakdown of all factors assessed.

A total of 17 resilience sub-dimensions and 39 related actions were identified across the six dimensions of resilience. Supplementary File 11 provides the themes (resilience sub-dimensions) identified in the survey, which were complemented and refined through focus groups discussions to develop the resilience tool. The list of resilience sub-dimensions and related actions are listed in Table 2 and presented briefly below.

**Table 2.**
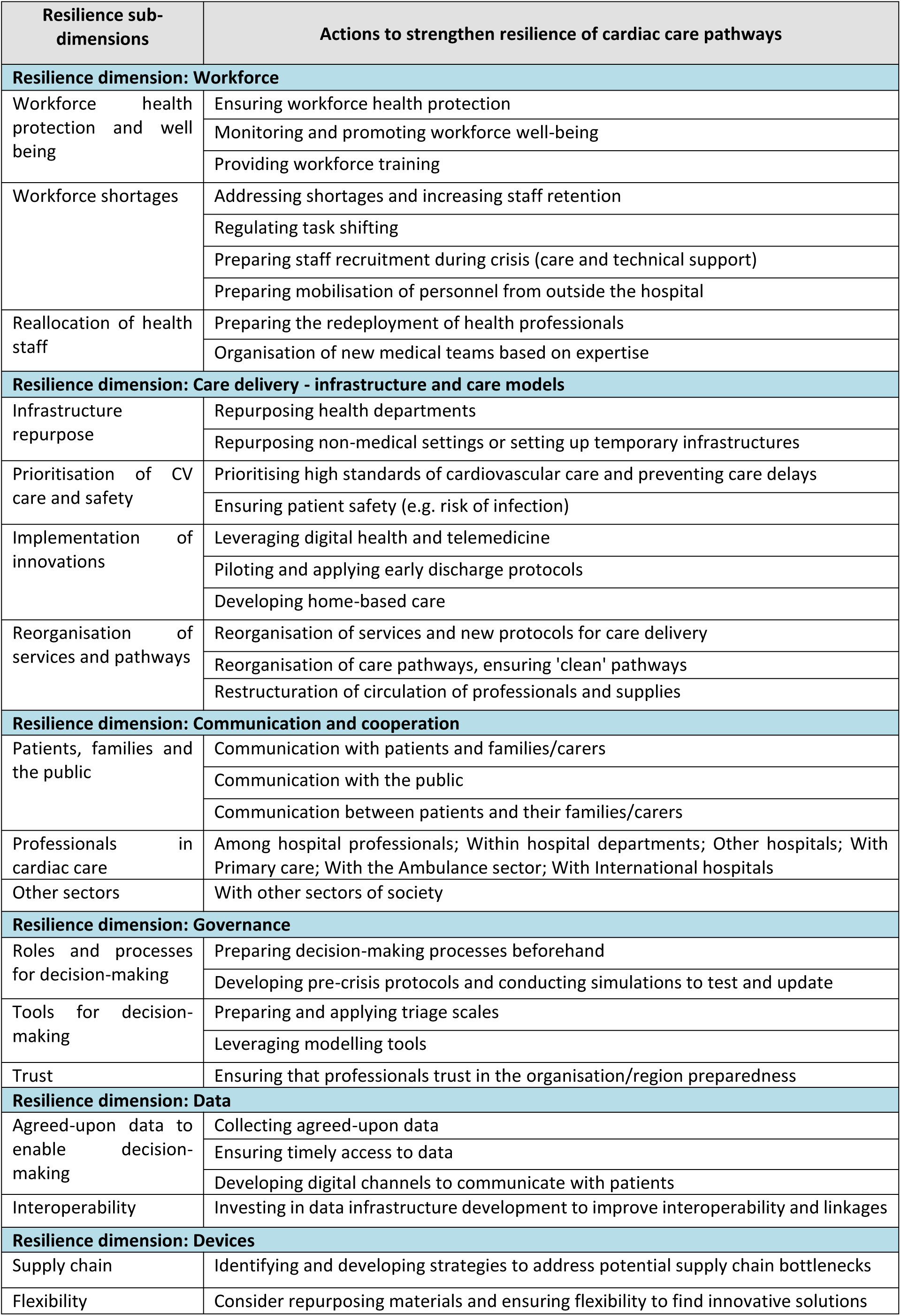
Summary of sub-dimensions of resilience and related actions to strengthen resilience.

| Resilience sub-dimensions | Actions to strengthen resilience of cardiac care pathways |
| --- | --- |
| **Resilience dimension: Workforce** |  |
| Workforce health protection and well-being | Ensuring workforce health protection |
|  | Monitoring and promoting workforce well-being |
|  | Providing workforce training |
| Workforce shortages | Addressing shortages and increasing staff retention |
|  | Regulating task shifting |
|  | Preparing staff recruitment during crisis (care and technical support) |
|  | Preparing mobilisation of personnel from outside the hospital |
| Reallocation of health staff | Preparing the redeployment of health professionals |
|  | Organisation of new medical teams based on expertise |
| **Resilience dimension: Care delivery - infrastructure and care models** |  |
| Infrastructure repurpose | Repurposing health departments |
|  | Repurposing non-medical settings or setting up temporary infrastructures |
| Prioritisation of CV care and safety | Prioritising high standards of cardiovascular care and preventing care delays |
|  | Ensuring patient safety (e.g. risk of infection) |
| Implementation of innovations | Leveraging digital health and telemedicine |
|  | Piloting and applying early discharge protocols |
|  | Developing home-based care |
| Reorganisation of services and pathways | Reorganisation of services and new protocols for care delivery |
|  | Reorganisation of care pathways, ensuring 'clean' pathways |
|  | Restructuration of circulation of professionals and supplies |
| **Resilience dimension: Communication and cooperation** |  |
| Patients, families and the public | Communication with patients and families/carers |
|  | Communication with the public |
|  | Communication between patients and their families/carers |
| Professionals in cardiac care | Among hospital professionals; Within hospital departments; Other hospitals; With Primary care; With the Ambulance sector; With International hospitals |
| Other sectors | With other sectors of society |
| **Resilience dimension: Governance** |  |
| Roles and processes for decision-making | Preparing decision-making processes beforehand |
|  | Developing pre-crisis protocols and conducting simulations to test and update |
| Tools for decision-making | Preparing and applying triage scales |
|  | Leveraging modelling tools |
| Trust | Ensuring that professionals trust in the organisation/region preparedness |
| **Resilience dimension: Data** |  |
| Agreed-upon data to enable decision-making | Collecting agreed-upon data |
|  | Ensuring timely access to data |
|  | Developing digital channels to communicate with patients |
| Interoperability | Investing in data infrastructure development to improve interoperability and linkages |
| **Resilience dimension: Devices** |  |
| Supply chain | Identifying and developing strategies to address potential supply chain bottlenecks |
| Flexibility | Consider repurposing materials and ensuring flexibility to find innovative solutions |

#### Workforce

Informants highlighted the critical importance of monitoring and promoting the health protection and well-being of healthcare professionals. Factors such as insufficient protective equipment, training, heightened risk of infection, long working hours, and frequent staff reallocation negatively affected staff well-being. While the ability to quickly mobilise professionals within and outside the hospital was considered key to addressing surges in care demand, this strategy poses challenges. One informant acknowledged that “[the willingness to be reallocated] was higher when we could keep the teams intact and move them together towards COVID care” (Head Nurse, Netherlands).

Addressing workforce shortages was also identified as a key factor to be able to absorb a crisis. Mobilising retired doctors and nurses was an aspect highlighted as helpful to prepare beforehand. As one informant stated: “ (…) [to be able to know] who does what and who doesn’t, what additional training is needed to be able to deploy and then arrange that very quickly.” (Representative of Emergency Medical Services, Europe).

#### Care delivery - infrastructure and care models

The ability to quickly repurpose and upgrade infrastructure and to establish temporary facilities in response to surges in care demand were recurrent themes. One informant mentioned: “we realised that converting a conventional room into an ICU was complicated, because of the issue of not being able to simultaneously put BiPAP or high flow. […]. […] So, I think that for the future, any unit, […] should be fully prepared to do this upgrade.” (Head of cardiology department, Spain).

Prioritising the continuity and quality of cardiac care during crises was also emphasized. Informants proposed the reorganisation of services and care pathways, including pre-defined ‘clean’ and ‘dirty’ pathways to sustain urgent and some elective care during crisis, as well as restructuring the movement of healthcare professionals, supplies, and equipment. Infrastructure should be prepared to accommodate this reorganisation, as one informant recognised: “For instance, we opened a brand-new hospital (…). However, despite being brand new, it did not account for the division between a dirty pathway and a clean pathway.” (Nurse, Italy). Implementing or scaling up innovations was also mentioned as contributing to strengthen the resilience of cardiac care pathways, such as home-based care models.

#### Communication and cooperation

Patient representatives and professionals highlighted the challenges related to the lack of information during the pandemic. Establishing official communication channels and providing targeted communication training were proposed as important strategies. As one informant underscored: “[…] communication with patients and the rest of the population is a protocol that fundamentally applies to any type of [crisis]” (Radiology technician, Italy).

Improving communication and cooperation within and beyond the health system was identified as a key priority. Within the healthcare system, collaboration among hospital professionals, across departments, with other hospitals, primary care providers, ambulance services, and international institutions should be in place before crises.

Furthermore, involving other societal sectors to ensure effective communication and coordinated responses was also mentioned, such as scientific societies and private sector health companies. One informant mentioned that “The other decision that was made from the point of view of the scientific society was not to stop scientific communications and scientific meetings. […] we learned a lot from each other […] thanks to the fact that scientific communication and the exchange of information did not stop. I think that these are things that the scientific community can and should do” (Cardiologist and representative of National scientific society, Netherlands).

#### Governance

Informants mentioned that governance structures should establish clear roles and decision-making processes in advance, supported by pre-crisis protocols and simulation exercises. As one informant highlighted: “we didn’t have pre-existing protocols on how to act in catastrophic or pandemic situations and I think this was something that we need to have for the future for a better response.” (General practitioner, Portugal).

Decision-making tools such as triage scales and modelling systems were identified as useful to support decision-making during crises. One participant underscored: “If you have a system, where all you do in a hospital is numbered: what is #1? What is #3? and what is #10? Then, if you have a crisis, […] if you have war, if you don’t have any money, then you could just cut off 10, 9, 8, 7, 6, 5, 4 and just do number 1, 2 and 3 or number 1 to 6. […] that is resilience. But that’s very difficult. […]” (National scientific organisation, Sweden). In Spain, a severity assessment scale was developed and validated to prioritise interhospital transfers.

Consistent public and patient involvement were also highlighted as a priority. Furthermore, building and maintaining trust among professionals in each organisations or region’s preparedness was emphasized to ensure a coordinated response during a crisis. As one informant acknowledged: “Regarding the delay in renewing the pandemic plan, I was involved in an exercise in 2012, […], at the Ministry of the Interior [where] we simulated a pandemic […] The problem was, in fact, […] translating those messages into everyday reality. When the COVID-19 pandemic began, none of what we had proposed was implemented. So that was the failure […]: the lack of belief in preparedness and organizing for such situations.” (Head of Intensive Care Cardiac Unit, Italy)

#### Data

Critical actions included improved digitalisation and health data infrastructure, particularly the collection of standardised data, timely access for decision-making, and the development of digital communication channels with patients. As one informant described: “So it’s always data. The big picture. What gives you control over it. […] where is there still capacity and how are we going to distribute it? I can predict that if we get another crisis like that, then everyone will start acting the same again and then it is extremely important to have insight into your exact numbers. Otherwise it will simply become chaos, just like it was in that first wave” (Representative of Emergency Medical Services, Europe).

Enhancing data infrastructure to support interoperability and system linkage was a recurrent theme to enable more coordinated responses. One informant highlighted: “Where I think the biggest problem lies is in getting a good view of what is happening outside the hospital - that data was very difficult to get insight into.” (Cardiologist, Netherlands)

#### Devices

To avoid disruptions in supply of products and medical devices during crises, identified actions were to focus on identifying potential bottlenecks and developing strategies to address them. Additionally, ensuring flexibility by repurposing materials and adopting innovative solutions to overcome equipment shortages was emphasised. As one informant explained: “And then devices that we didn’t have, not only ventilators, but positive pressure devices […] We had to adapt a little that we had at our disposal, with the help of an engineer […] to free up a resource in the ICU.” (Emergency department physician, Spain).

### Structure of the Resilience Assessment Tool

Considering these data collected and based on discussion among the RESIL-Card Consortium, it was agreed that the tool would be designed to enhance the resilience to face various types of crises, targeting healthcare professionals in the hospital setting, while ensuring involvement of all professionals working with them and interaction with patients and the general population. Furthermore, it was agreed that the tool would include: a) an introductory component to highlight the importance of prioritising emergent cardiac care, b) the definition of resilience explicitly and repetitively in the tool, c) clear guidance for target users on how to use the tool, d) a component of social interaction to ensure that professionals from the same context would meet to assess their preparedness level and discuss actions to improve it, through a preparedness checklist, which would generate a preparedness visual heat map.

The tool comprises four key components (Figure 4) and is available at https://doi.org/10.5281/zenodo.16994435. First, users are guided to thoroughly describe the cardiac care pathway within their specific context. Second, users are prompted to identify the key actors who should collaboratively use the tool to conduct the preparedness exercise. Third, a group exercise involving a self-assessment checklist is provided. Upon completion, the checklist generates a visual heat map that highlights potential areas for improvement in each context. The fourth component is a toolkit with links to additional resources, including a list of good practices and literature compiled by the RESIL-Card Consortium, alongside a set of recommendations to support users in designing their actions.

**Figure 4.**
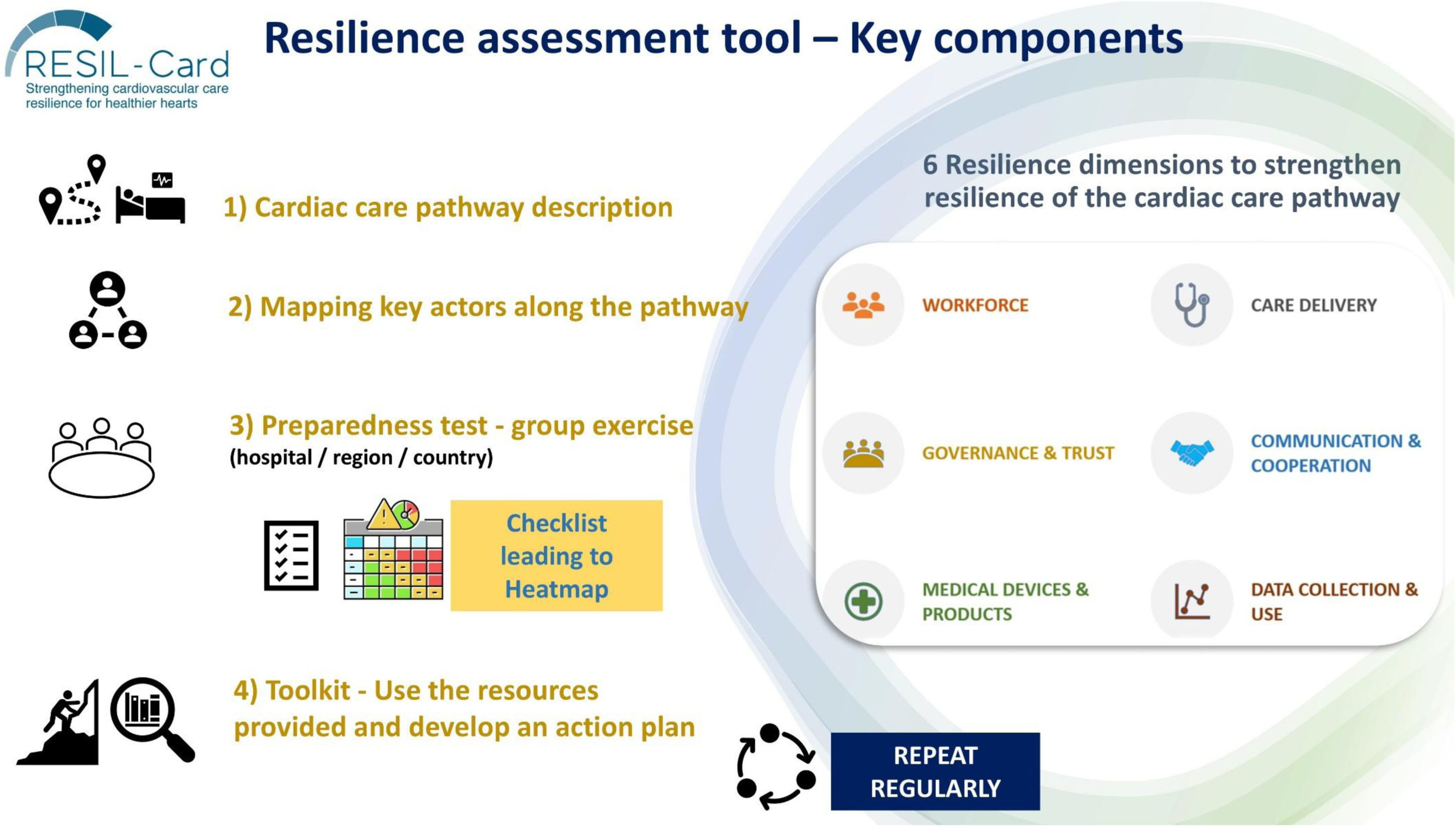
Structure of the Resilience Assessment Tool.

## 4. DISCUSSION

This study aimed to develop a structured and context-sensitive tool to assess and strengthen the resilience of cardiac care pathways. Using a sequential mixed-methods design, we first identified key challenges experienced during the COVID-19 pandemic and then translated these into a tailored self-assessment tool. The study identified six dimensions of health system resilience from the literature as relevant to cardiac care pathways: (a) workforce, (b) organisation of care delivery, (c) governance and trust, (d) communication and cooperation, (e) medical devices and products, and (f) data collection and use. A survey among cardiology healthcare professionals revealed that the most significant challenges to ensure cardiac care continuity during the COVID-19 pandemic were related to the first two dimensions, notably shortages of staff and infrastructure capacity. The factors considered most crucial to maintain access to cardiac care during crises were investing in the workforce’s health protection and strengthening communication with patients. A total of 17 resilience sub-dimensions were identified, and 39 concrete actions were recommended to strengthen the resilience of cardiac care pathways, which culminated in the development of the resilience assessment tool.

Our study showed that addressing workforce shortages, investing in their health protection, and ensuring the ability to mobilise health workers should be prioritised to prepare for future crises, which is in line with previous studies. A survey to around 600 interventional cardiologists conducted in April 2020 reported that the availability of personnel in cardiac catheterisation laboratories was affected by COVID-19 infections, quarantine, or staff reallocation [26]. Suboptimal availability of personal protective equipment was also reported [26]. The WHO/Europe framework for ‘resilient and sustainable health systems’ adopted in 2024 includes workforce training and support one of its four pillars, alongside models of care delivery, universal health coverage, and person-centred care [27]. The WHO study on ‘Health systems resilience during COVID-19’ likewise emphasised the mobilisation and support, scaling up capacity, and investing in training, flexibility and workforce’s health [16].

Given the substantial burden of cardiac diseases and the importance of timely access to care on patient outcomes, maintaining quality standards is particularly critical in this clinical area. With respect to infrastructure shortages during crises, our results identified crucial aspects to ensure resilient cardiac care pathways: the ability to swiftly repurpose and upgrade medical infrastructure, to uphold high standards of cardiovascular care, notably leveraging innovative care models and flexible care pathways. A study in Dutch hospitals showed that flexibility measures related to building design were the most practical solutions to implement during COVID-19, namely creating triage areas and separate clean and dirty routes [28]. However, this study also highlights that flexibility in facility design is limited by the staff’s adaptability and availability [28], underscoring that a combination of factors needs to be addressed to improve crisis resilience.

In Organisation for Economic Co-operation and Development (OECD) member countries, 13 out of 19 countries reported expanding their hospital networks, underscoring that collaboration was an important factor to foster resilience [15], which is in line with the present results. Governance aspects were reported in previous studies to be an important, nonetheless frequently overlooked, area of resilience [29]. In our study, establishing clear roles and decision-making processes in advance, supported by pre-crisis protocols and simulation exercises were also considered key elements of preparedness.

In agreement with our results regarding the need to invest in health data infrastructure to support decision-making, an OECD survey conducted in 2019-2020 showed that only 14 OECD countries had the ability to link data across different health system settings [15]. After the COVID-19 pandemic, data linkage and data coverage have improved across OECD countries, although data quality and data interoperability still need improvement [30].

Supply chain problems were reported by 16 out of 24 OECD countries at the early stages of the pandemic [15]. Proposed strategies are regulatory changes, internal production and supply chain diversification [15]. This corroborates our findings, underscoring the importance of identifying and addressing potential supply chains bottlenecks to enhance preparedness.

In November 2024, the Council of the European Union (EU) has approved a set of conclusions related to cardiovascular health in the EU [31], where this clinical area was considered a key priority. Furthermore, in May 2025, the WHO Pandemic Agreement was formally adopted by the 78^th^ World Health Assembly. The European commitment on improving the prevention and management of cardiovascular diseases, alongside the global focus on enhancing pandemic preparedness focusing in strengthening international coordination underscores the pressing need to empower professionals and policymakers on this topic.

At the policy level, the findings from this study may inform the development and implementation of a European Cardiovascular health plan. Specifically, the six resilience dimensions, their corresponding sub-dimensions and the outlined actions could contribute to define policy priorities aimed at enhancing the resilience of cardiac care pathways. At organisational level, these findings may support healthcare professionals in identifying key priorities in their context, such as developing decision-making processes, pre-crisis protocols, conducting simulations, and strengthening cooperation with other hospitals and with primary care providers. Healthcare professionals should play a key role in implementing the actions outlined in the resilience tool, ensuring the inclusion of patient representatives and scientific societies.

### Strengths and limitations

While a growing body of literature provides guidance on enhancing health system resilience [12, 15, 16], this study has the strength of focusing specifically on cardiac care delivery, building on lessons from the COVID-19 crisis and addressing the lack of standardised tools tailored to cardiac care pathways. Following a sequential mixed-methods approach, key challenges and strategies were identified in this study, from the perspective of stakeholders across health system levels across various European countries, facilitating cross-learning among organisations and countries. The regular involvement of the RESIL-Card Consortium and its Advisory Board allowed to integrate various perspectives in the tool development, enhancing the applicability of our findings and the fitness-for-use of the tool. Cross-learning across countries was previously acknowledged as a lever to enhance resilience [29].

This study has some limitations. We obtained a low survey response rate, with an overrepresentation of Italian and Spanish cardiologists. Nonetheless, Italy was the first European country severely impacted by the COVID-19 crisis in Europe [32], being, therefore, a very relevant country to collect insights from the stakeholders’ experience. Furthermore, the sequential mixed-method design allowed to discuss and complement the quantitative survey’s findings in the three focus groups with stakeholders from other European countries. The survey’s low response rate limits the ability to inform how each resilience dimension should be prioritised to enhance the resilience of cardiac care pathways. Nonetheless, informed by the survey and focus groups’ findings, the resilience assessment tool was designed to support users in identifying the weaknesses and strengths in their own context through a preparedness checklist leading to a heat map. Furthermore, the tool provides recommendations and resources to support end-users developing an action plan applicable to their own context.

## 5. CONCLUSION

Leveraging the COVID-19 experiences to develop actionable insights is essential for building more resilient healthcare systems. This study aimed to develop a structured and context-sensitive tool to assess and strengthen preparedness and response to a future crisis affecting cardiac care delivery in Europe. The resulting resilience assessment tool provides a step-by-step guidance to resilience building across six resilience dimensions, 17 resilience sub-dimensions, providing concrete actionable recommendations, while fostering collaboration among stakeholders in the cardiology ecosystem.

The survey’s low response rate limited the ability to inform on the prioritisation of each resilience dimension, nonetheless the sequential mixed-method design allowed to discuss and complement the findings through five multi-stakeholder focus groups, followed by a collaborative multi-stakeholder development process. The pilot phase, which has started in early-2025, will ensure that the tool meets the needs of end-users, paving the way for wider implementation.

The tool may support policymakers and healthcare professionals in identifying and prioritising context-specific vulnerabilities and improvement priorities to strengthen the cardiac care pathway. Ultimately, the tool may advance the ability of health systems to cope with shocks and ensure high-quality cardiac care delivery before, during and after crises. Future research could test and adapt these findings to other clinical areas and geographies, contributing to develop more efficient, equitable and patient-centred health systems across Europe and globally.

## Supporting information

Completed checklist for mixed methods research manuscript preparation and review

Cognitive testing interviews for survey development

RESIL-Card survey

Organisations and initiatives to raise awareness to the RESIL-Card questionnaire

Focus groups - Breakdown of total number of invitations and participation

Focus groups: Detailed breakdown of informants

Survey results, Section1: Challenges before the COVID-19 pandemic

Survey results, Section2: Nature of disruptions during the COVID-19 pandemic and the practices and innovations implemented

Survey results, Section3: Lessons learned after the COVID-19 pandemic

Themes identified to develop the resilience tool, per resilience dimension

## Data Availability

The datasets generated and analysed during the current study are available from the Zenodo repository, 10.5281/zenodo.16993253.

https://zenodo.org/records/16993254

## ACKNOWLEDGMENTS

We sincerely thank all professionals who generously contributed with their time and expertise through cognitive testing of the survey, completing survey responses, and participating in the focus groups. We are grateful for the support of ‘Global Heart Hub’, whose collaboration was essential in ensuring the inclusion of the patient perspective throughout all the phases of this work. We would like to express our sincere gratitude to Caius Merșa and Marcin Ruciński for their valuable contribution as patient participant in the focus groups, which were instrumental to incorporate the patient perspective in this work. We are also grateful to the RESIL-Card Consortium for their valuable input throughout the project. We extend our appreciation to the RESIL-Card’ Partnering organisations and Advisory Board for their close involvement and contributions.

Preliminary results of this work have been presented in a scientific conference for Interventional Cardiologists, EuroPCR, in May 2025.

## AUTHOR CONTRIBUTIONS

NK, ASVC, OBF, DK conceptualised the study. All authors (ASVC, OBF, JJP, JM, ASE, WW, NK, DK) contributed to data collection. ASVC performed the data analysis and prepared the initial manuscript draft. All authors (ASVC, OBF, JJP, JM, ASE, WW, NK, DK) reviewed the manuscript, provided critical feedback, and approved the final version for publication.

## PATIENT AND PUBLIC INVOLVEMENT

Patients and/or patient representatives were involved in the conduct, reporting, and dissemination of this research. Further details are provided in the Methods section.

## PATIENT CONSENT FOR PUBLICATION

Not applicable.

## ETHICS APPROVAL

This study was approved by the non-WMO Committee of the Medical Ethics Review Committee of Amsterdam University Medical Centers: committee’s reference number 2024.0020, on the 14^th^ Feb 2024. This committee is authorised by the board of directors of Amsterdam UMC and supervised by the Medical Ethics Review committee. Participants in the study provided written informed consent during the recruitment phase and expressed their consent verbally at the start of the focus groups. Survey and focus group data has been anonymised and confidentiality was assured by referring to informants by country name.

## AVAILABILITY OF DATA AND MATERIALS

The datasets generated and analysed during the current study are available from the Zenodo repository, https://zenodo.org/records/16993254.

