## Supplementary material for "Development of a resilience assessment tool for cardiac care pathways in Europe: A mixed-methods study": Completed checklist for mixed methods research manuscript preparation and review

### Supplementary File 1 - Completed checklist for mixed methods research manuscript preparation and review (Lee et al, 2022)*

| **Checklist item** | **Reported on Page or Additional file #** |
| --- | --- |
| Rational and description of MMRdesign | |
| Provide a clear statement of the study purpose | Last paragraph of the Introduction section. |
| Explicitly describe the MMR design in accordance with Creswell’s (2015) typology and use a diagram to illustrate the relationship and sequence of qualitative and quantitative research components | First paragraph of Methods section and Figure 1. |
| Justify why the MMR design is appropriate for meeting the study purpose | First paragraph of Methods section. |
| Transparency in describing method details | |
| Describe the study population(s) and sample(s; e.g., who, what, how many) | Methods section, Data collection sub-section, in the first paragraph of Phase 1 and Phase 2. |
| Describe the sampling procedures (including inclusion and exclusion criteria, recruitment) | Methods section, Data collection sub-section, in the first paragraph of Phase 1 and first two paragraphs of Phase 2. |
| Describe qualitative data collection processes (how often data were collected, who collected the data, what kind of data collection instruments were used, how data were recorded—e.g., notes, transcripts) | Methods section, Data collection sub-section, third paragraph of Phase 1 and of Phase 2. |
| Describe quantitative data collection processes (how often data were collected, who collected the data, what kind of data collection instruments were used measurements, validity/reliability) | Methods section, Data collection sub-section, third paragraph of Phase 1. |
| Describe qualitative data analysis processes (coding, single or multiple coders, replication logic, credibility) | Methods section, Data analysis sub-section, under ‘Focus groups data’ |
| Describe quantitative data analysis procedures (missing data and how they are handled, statistical tests used) | Methods section, Data analysis sub-section, under ‘Survey data’ |
| Integration of qualitative and quantitative research components | |
| Interpret qualitative analysis results with appropriate quotes if necessary | Results section, sub-section ‘Actions recommended to strengthen the resilience of cardiac care pathways’, under ‘workforce’, ‘care delivery’, ‘communication and cooperation’, ‘governance’, ‘data’, ‘devices’. |
| Interpret quantitative analysis results in consideration of statistical significance, selection bias, and threats to validity | Not applicable. |
| Compare qualitative and quantitative results | Methods section, Data analysis sub-section, under ‘Synthesis of data to develop the resilience assessment tool’. |
| Address divergencies and inconsistencies between qualitative and quantitative results | Methods section, Data analysis sub-section, under ‘Synthesis of data to develop the resilience assessment tool’. |
