## Supplementary material for "Development of a resilience assessment tool for cardiac care pathways in Europe: A mixed-methods study": Cognitive testing interviews for survey development

### Supplemental File 2 - Cognitive testing interviews for survey development: interview date, participants, questions addressed, and changes to the survey

| **Speciality, location, specialisation/role, and gender of the participants** | **Survey questions discussed** | **Interview date and number of people interviewed** |
| --- | --- | --- |
| **1- Senior cardiologist consultant**  Location: University Hospital, Belgium  Specialisation: Cardiology and ischemic heart disease  Gender: Man | Section 1: questions 1-53  Section 2: sub-sections a) and b)  Final questions about Resilience | 16th April 2024  2 people in the same interview |
| **2- Head nurse of cardiology ward**  Location: University Hospital, Belgium  Gender: Woman | Section 1: questions 1-53  Section 2: sub-sections a) and b)  Final questions about Resilience | 16th April 2024  2 people in the same interview |
| **3- Cardio-vascular surgeon**  Location: University Hospital, Belgium  Role: Chief Information Officer  Gender: Man | Section 2  Section 3 | 17th April 2024  Individual interview |
| **The draft was adjusted after these 2 interviews (18^th^ April 2024),** based on the most relevant comments and the ones possible to address in a short period of time, specifically:   - examples of diseases where telemedicine was used were added; - options were added in some questions, such as the ‘patient involvement’ and ‘chief medical officer’ in decision-making processes - some questions were written differently to make them clearer; - options were re-ordered to what was making more sense to respondents; - description in the numbers was added in the ranking questions ; - “I don’t know” option was added in some questions. | | |
| **4- Senior cardiologist**  Location: University hospital, Belgium  Specialties: Interventional Cardiology and Hypertension  Gender: Man | Complete survey | 19^th^ Apr 2024  Individual interview |
| **5- Cardiology business unit manager**  Location: University hospital, Belgium  Gender: Woman | Section 2: questions 29 – 45  Final questions about Resilience | 19^th^ Apr 2024  2 people in the same interview |
| **6- Head Nurse Cardiac Care Unit**  Location: University hospital, Belgium  Gender: Man | Section 2: questions 29 – 45  Final questions about Resilience | 19^th^ Apr 2024  2 people in the same interview |

After cognitive testing, the following adjustments were made to the survey:

- section 1 - before the pandemic: the cognitive testing showed it was a very long section and respondents were already thinking about the pandemic, which made it hard to answer to the questions before the pandemic. All questions were removed, and one new question was added, asking about the main challenges present before the pandemic in each respondent context;
- section 2 - during the pandemic: some questions were rephrased to make them clearer; the ranking of some questions was reordered and made homogeneous across the survey; "not applicable” and “I don’t know” options were added to some questions;
- section 3 - after the pandemic: the sections presented in this section were removed, and 2 new sections were created, so that the respondents, after reflecting on the experience of the pandemic, were immediately asked their opinion about ranking components most useful to have in a resilience tool; the open-questions were grouped, as they were found to be hard to reply in cognitive testing. The questions about workforce, data, and care delivery after the pandemic were moved to the end of this section, to reflect on the present situation.
