## Supplementary material for "Development of a resilience assessment tool for cardiac care pathways in Europe: A mixed-methods study": RESIL-Card survey

### Supplementary file 3 - RESIL-Card survey

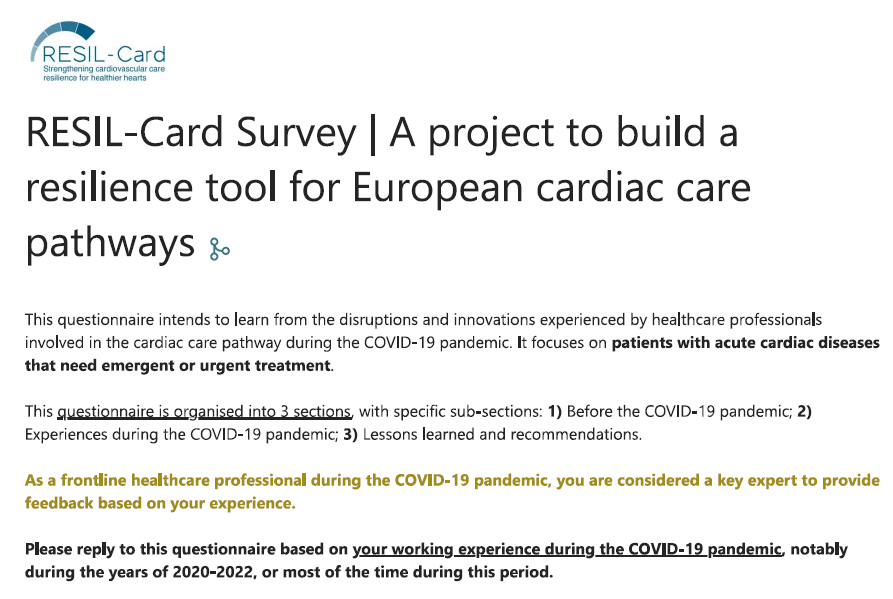

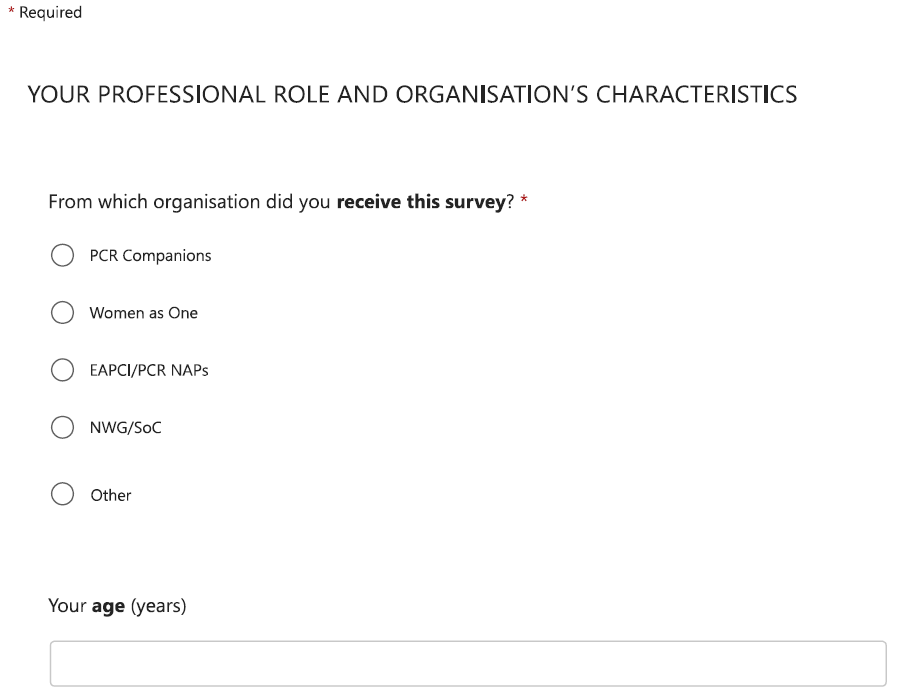

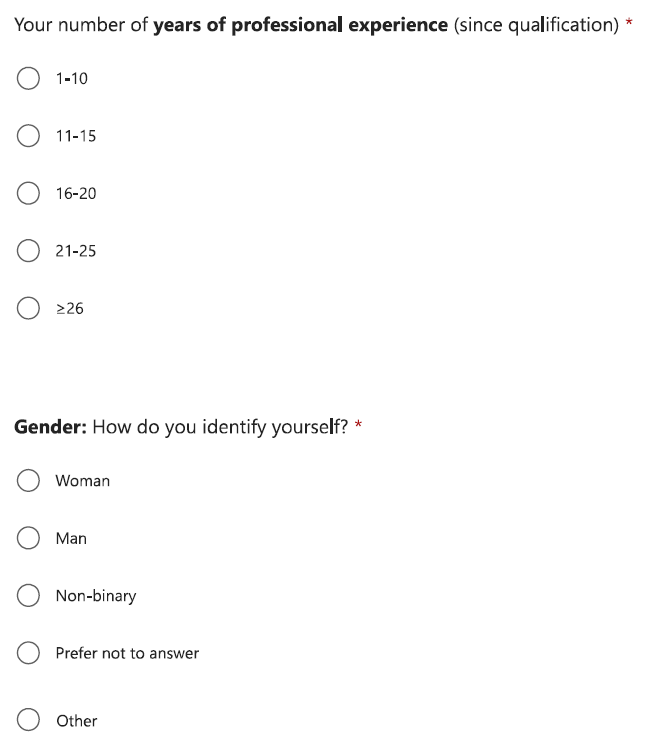

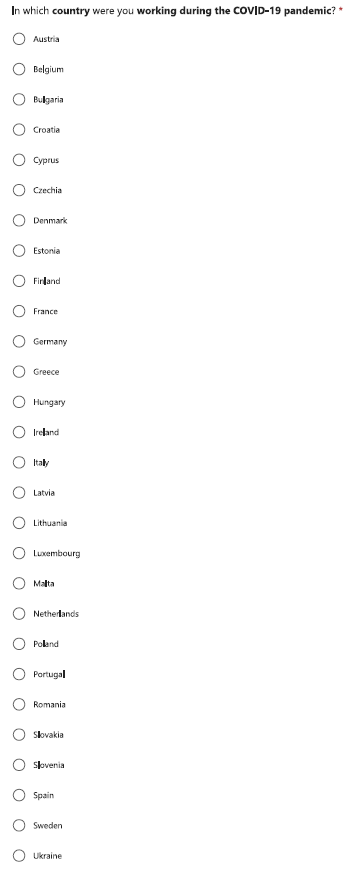

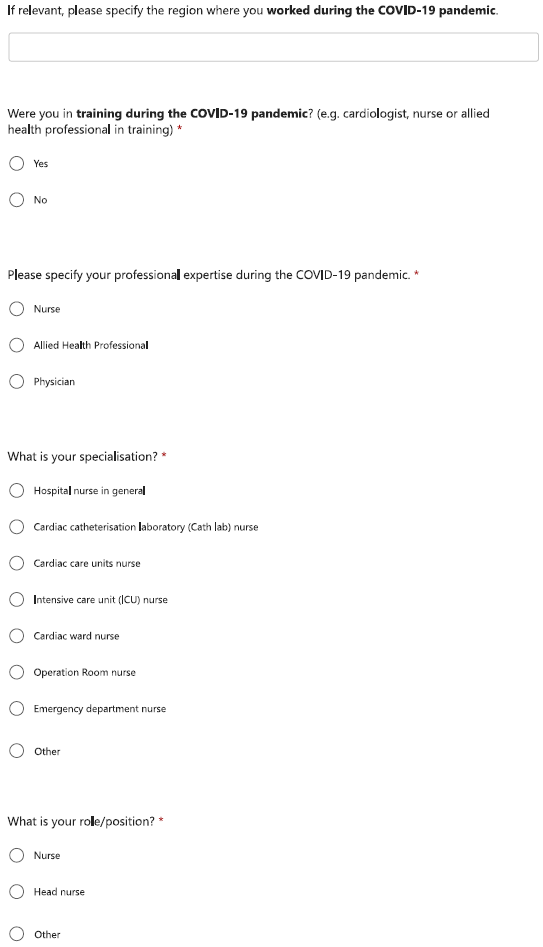

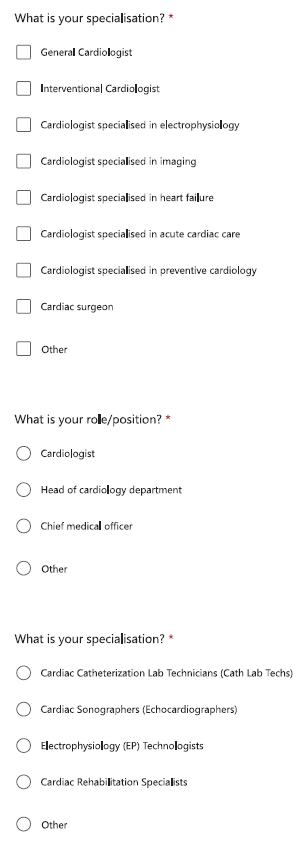

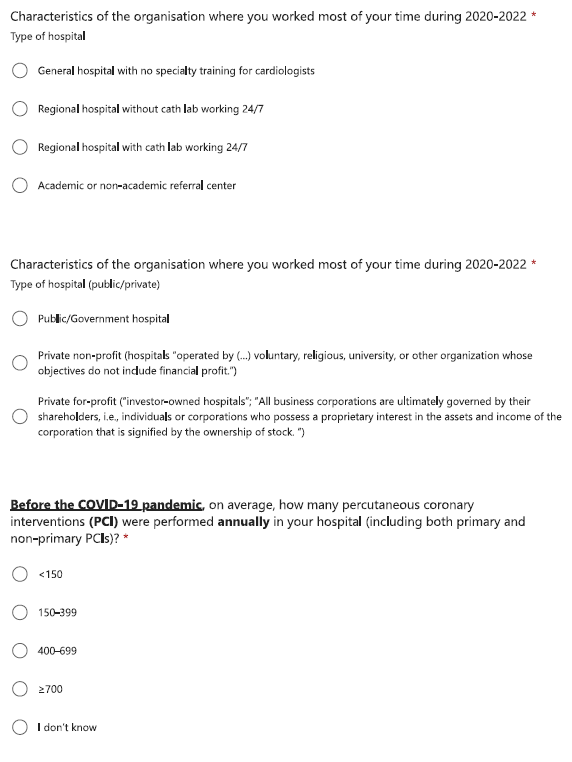

#### Section 1 - before the COVID-19 pandemic

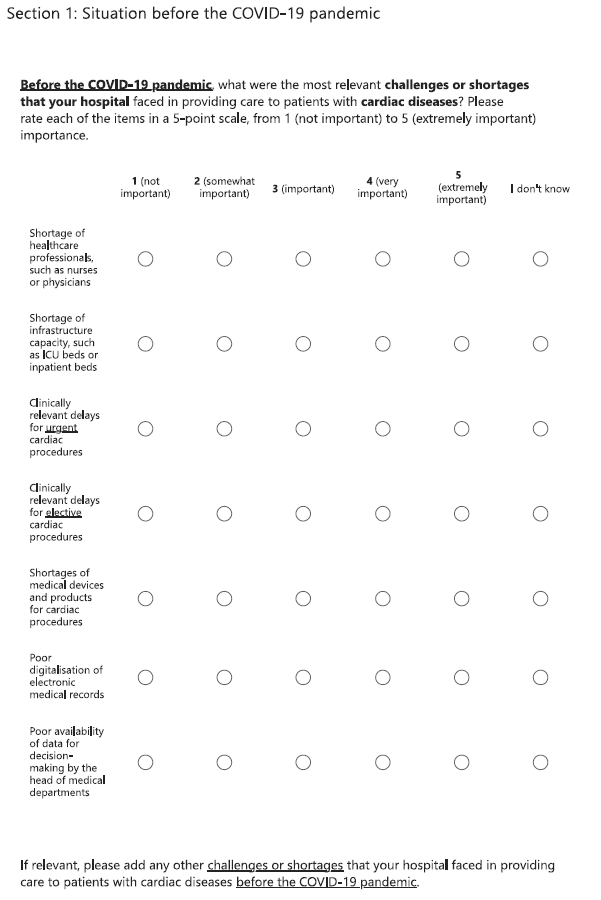

#### Section 2 - during the COVID-19 pandemic

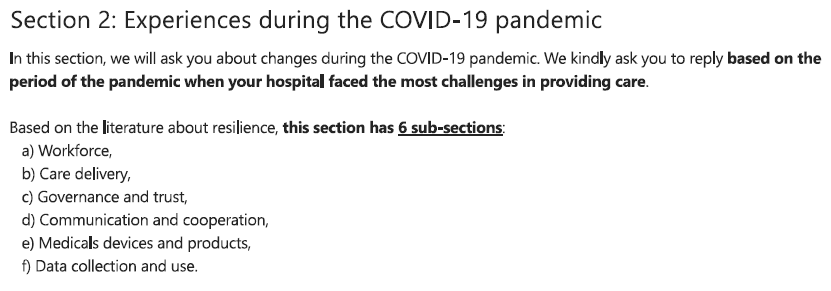

#### a) Workforce

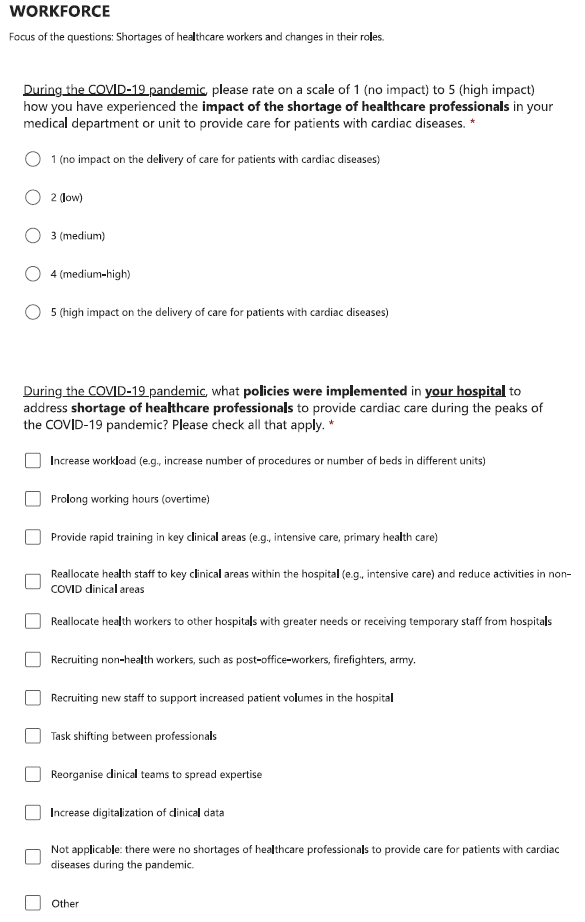

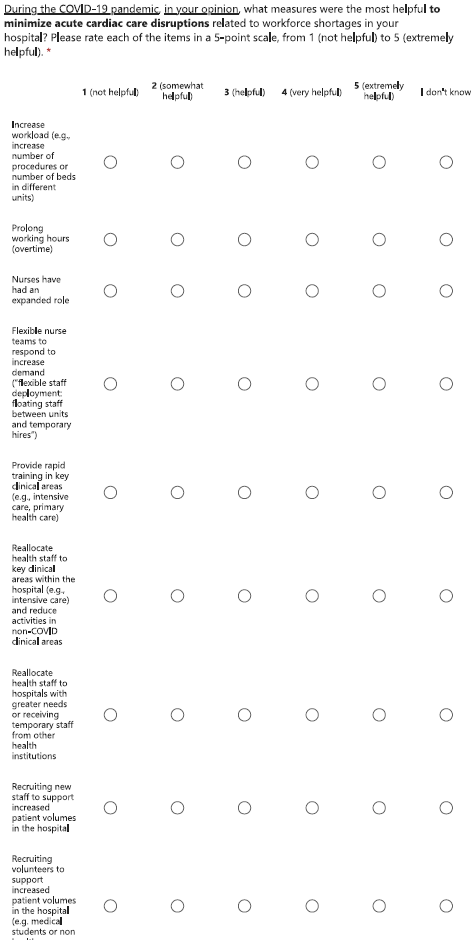

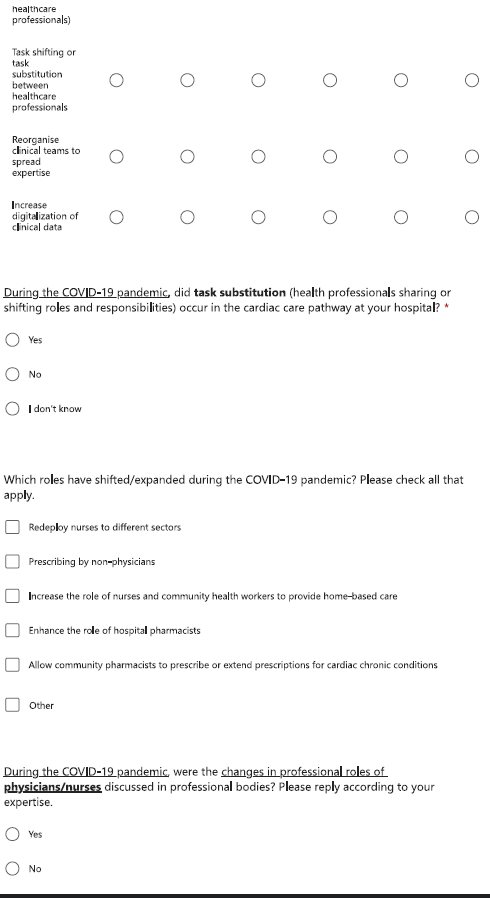

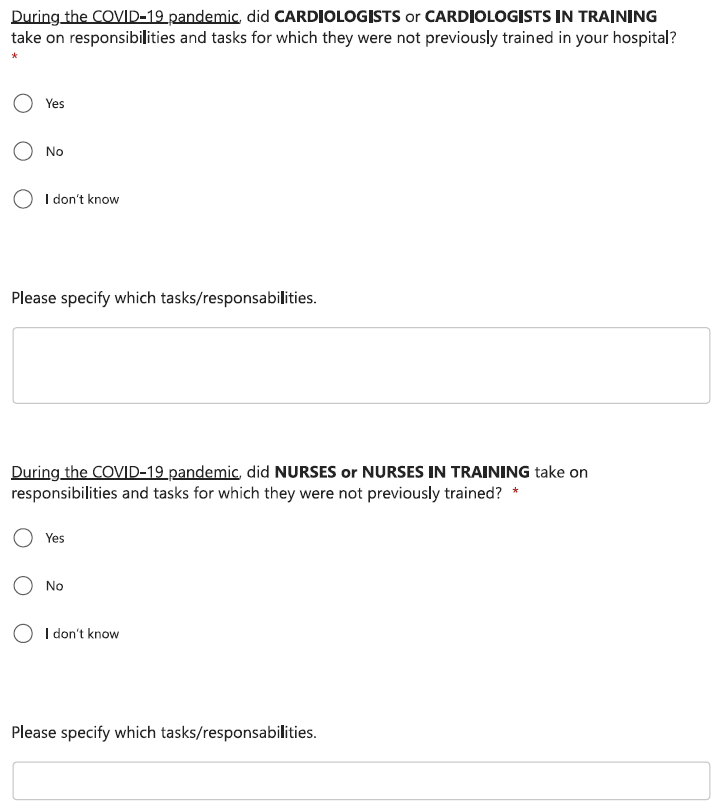

#### b) Care delivery

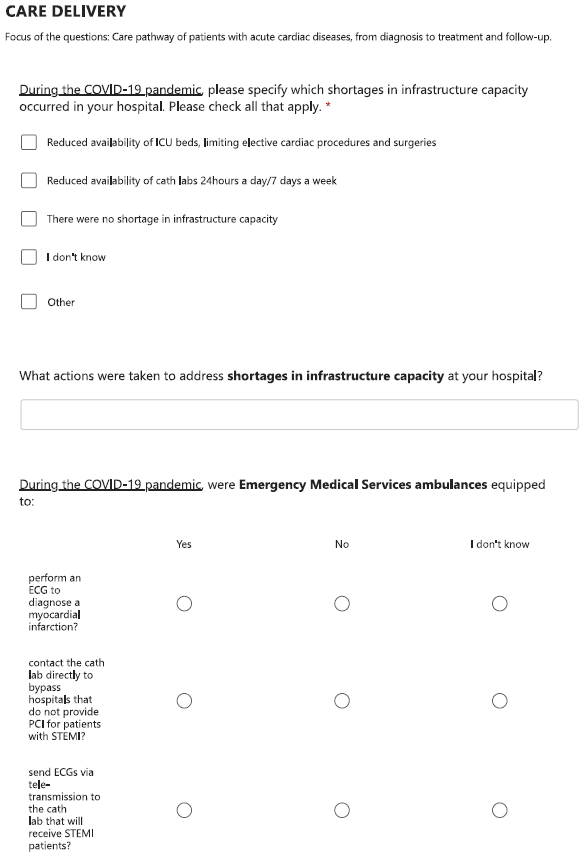

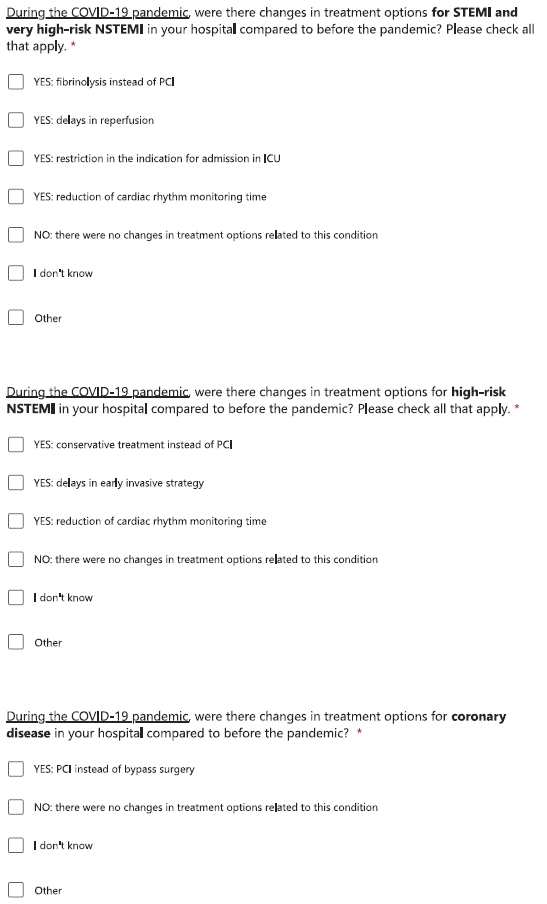

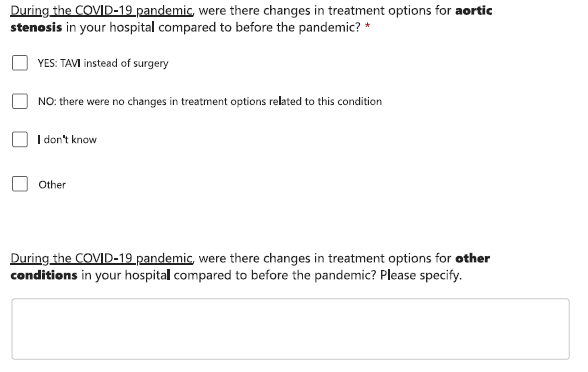

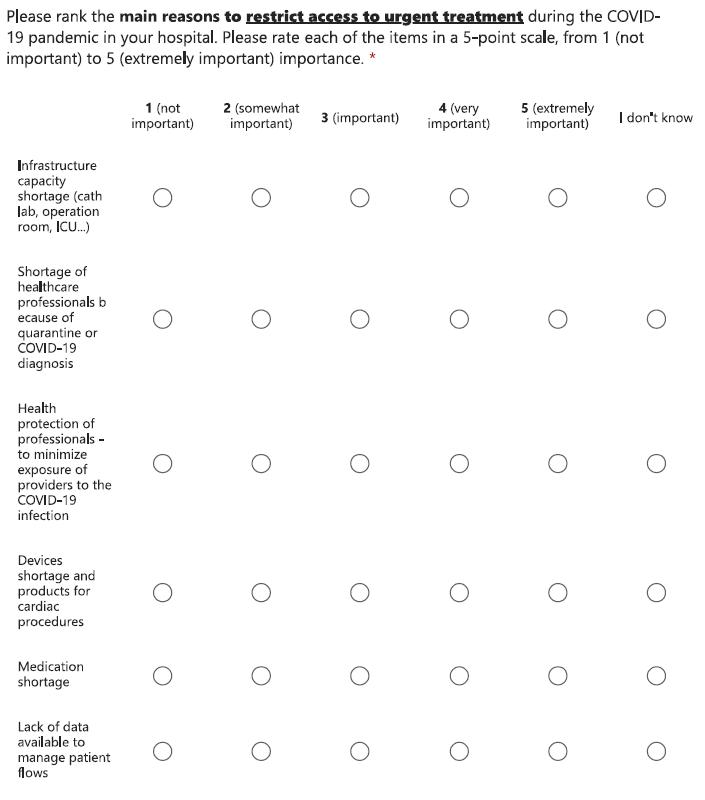

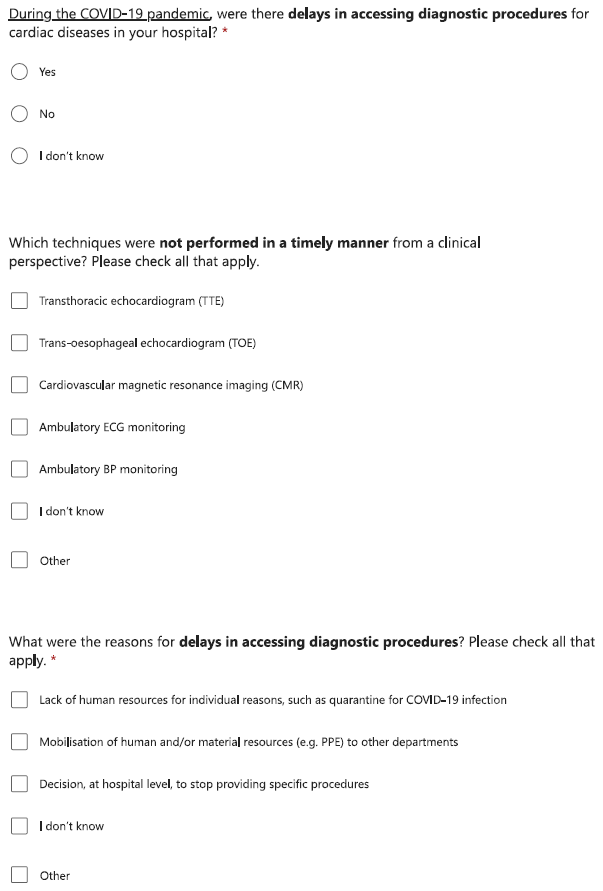

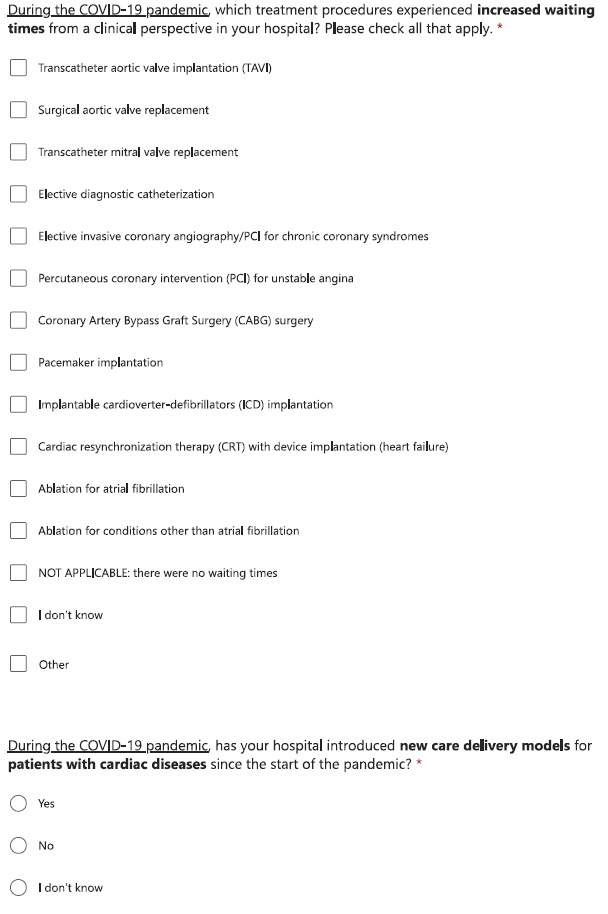

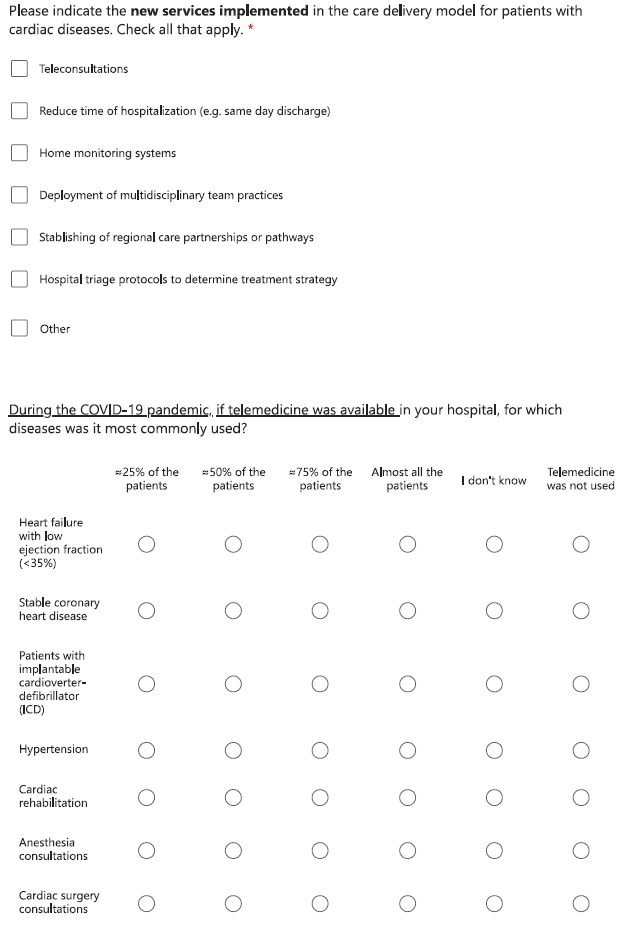

#### c) Governance and trust

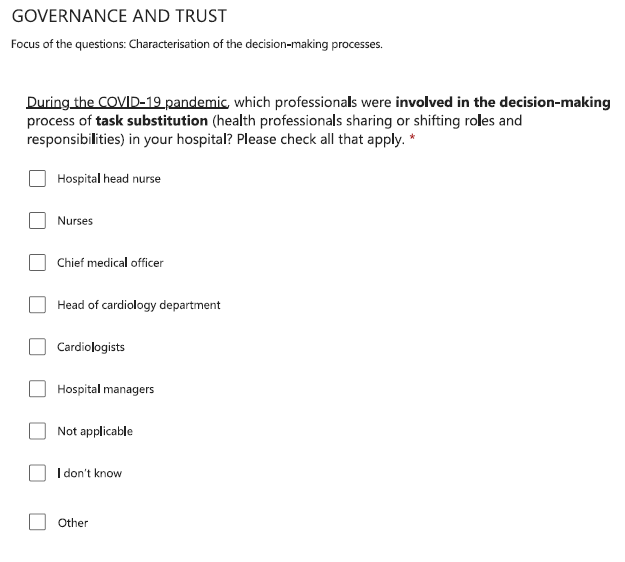

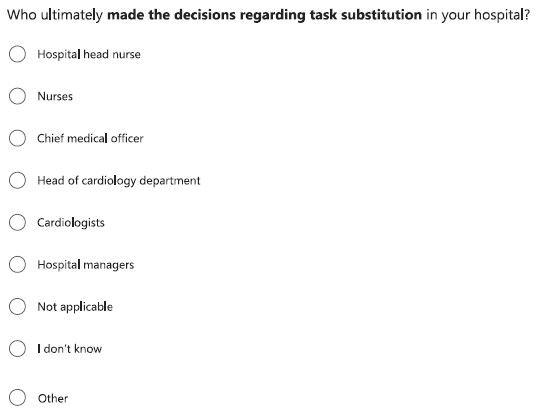

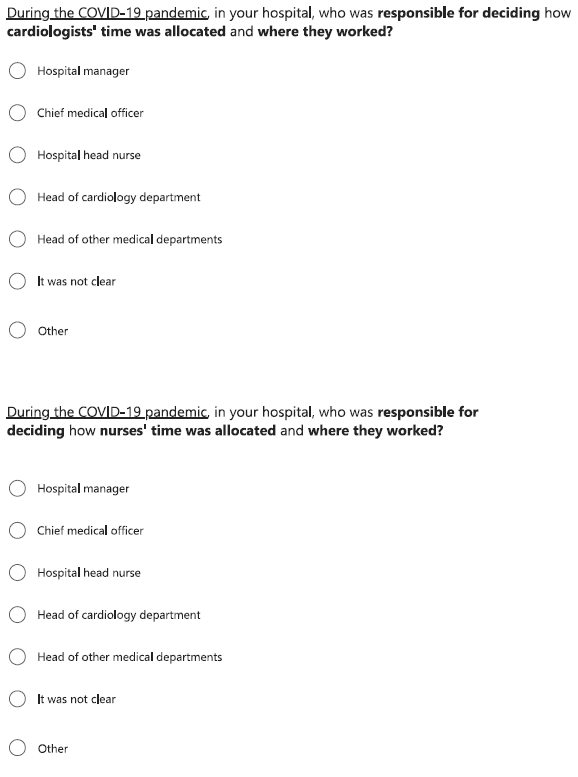

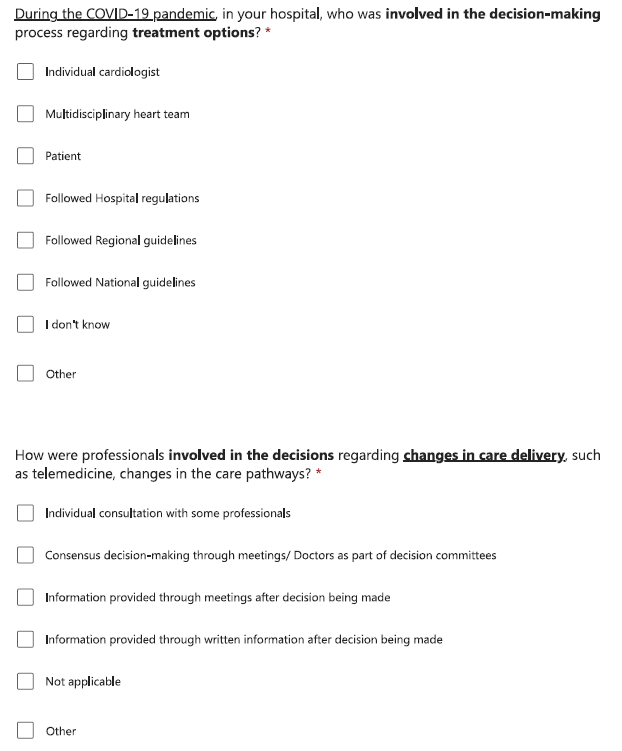

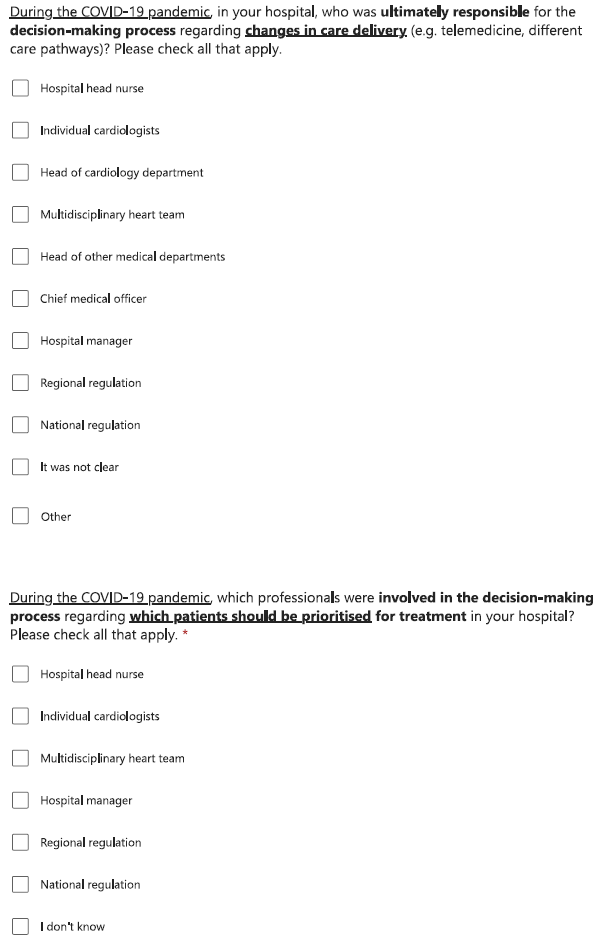

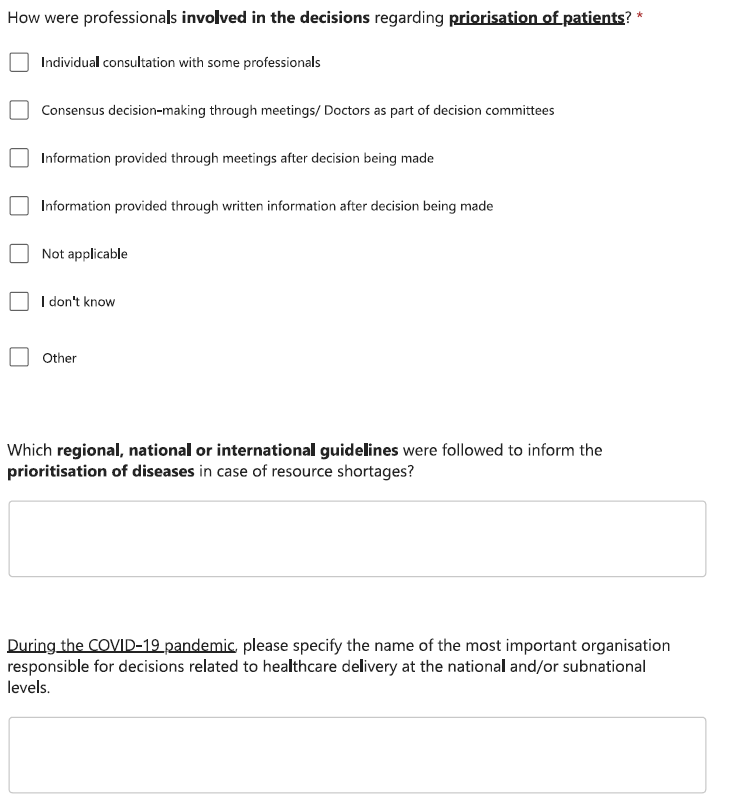

#### d) Communication and cooperation

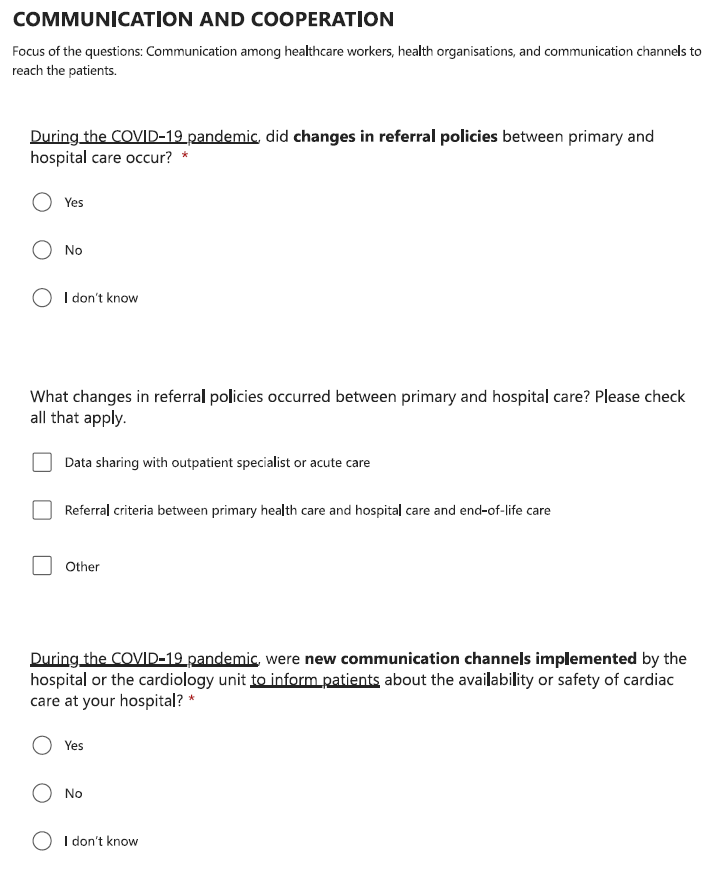

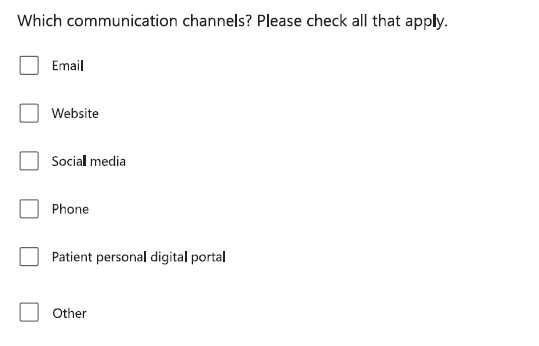

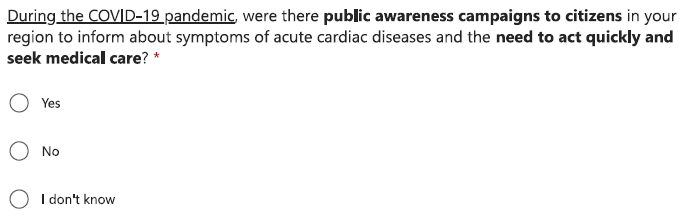

#### e) Medicals devices and products

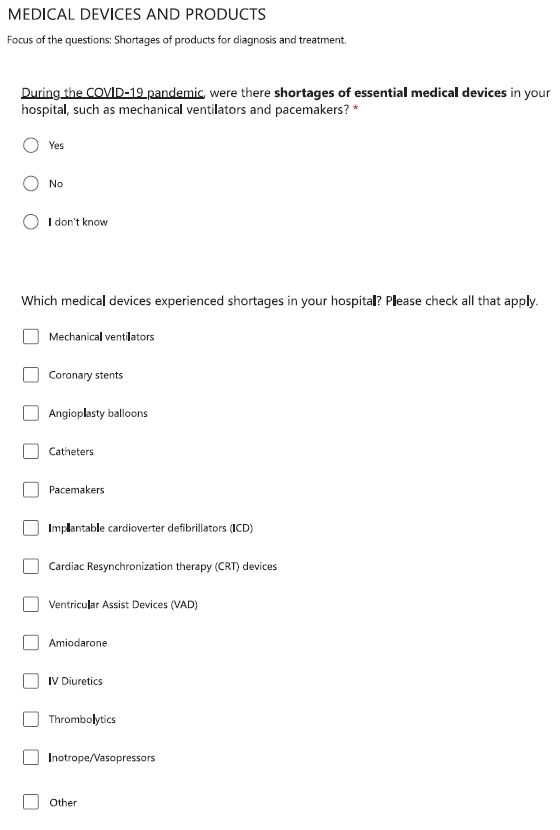

#### f) Data collection and use

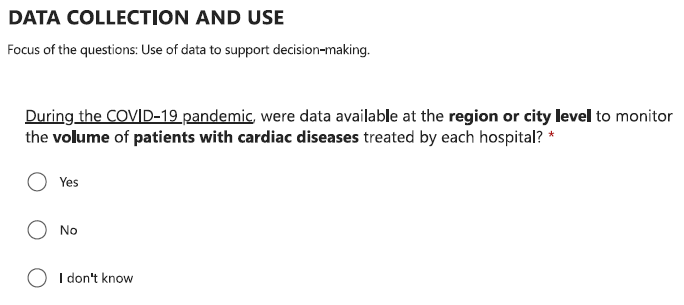

#### Section 3 – after the COVID-19 pandemic

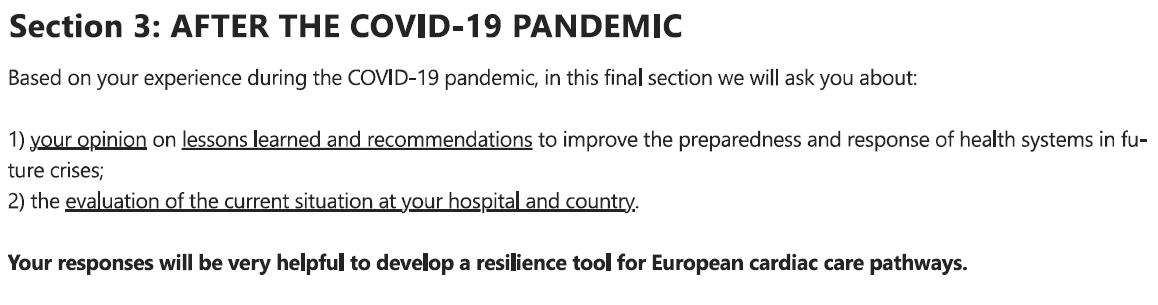

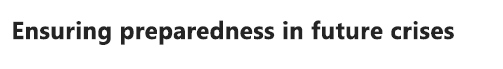

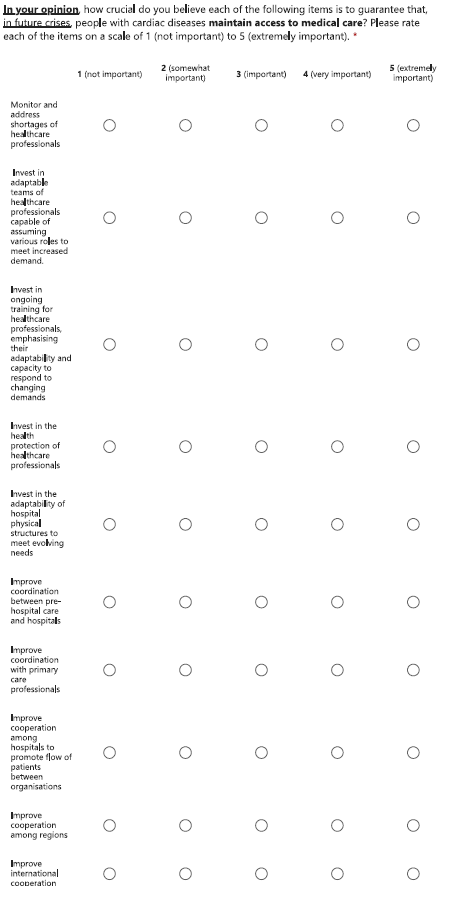

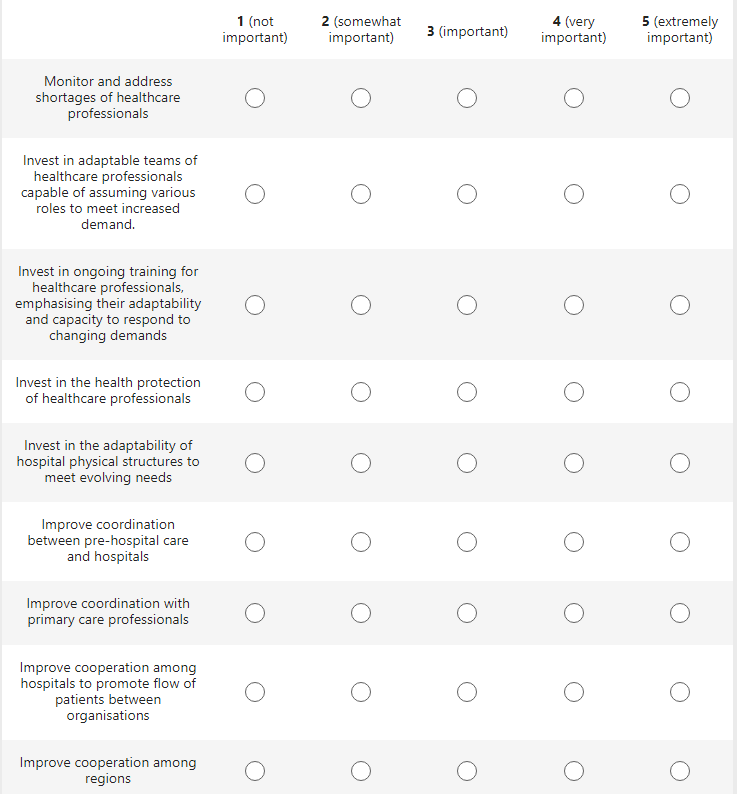

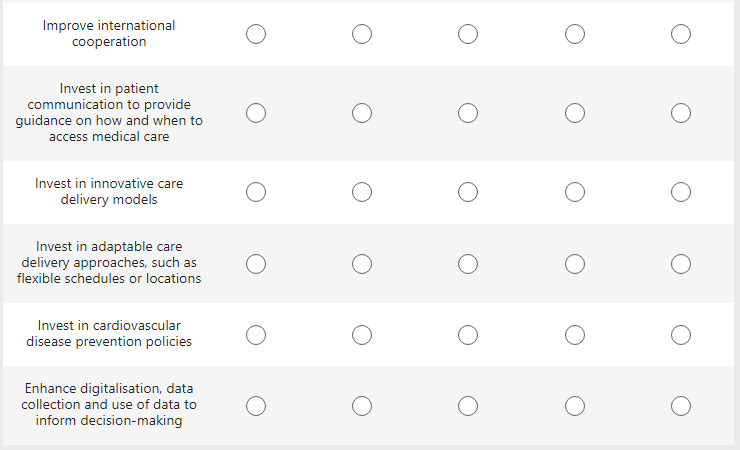

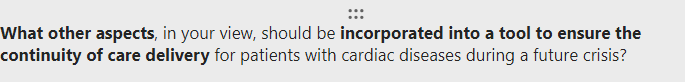

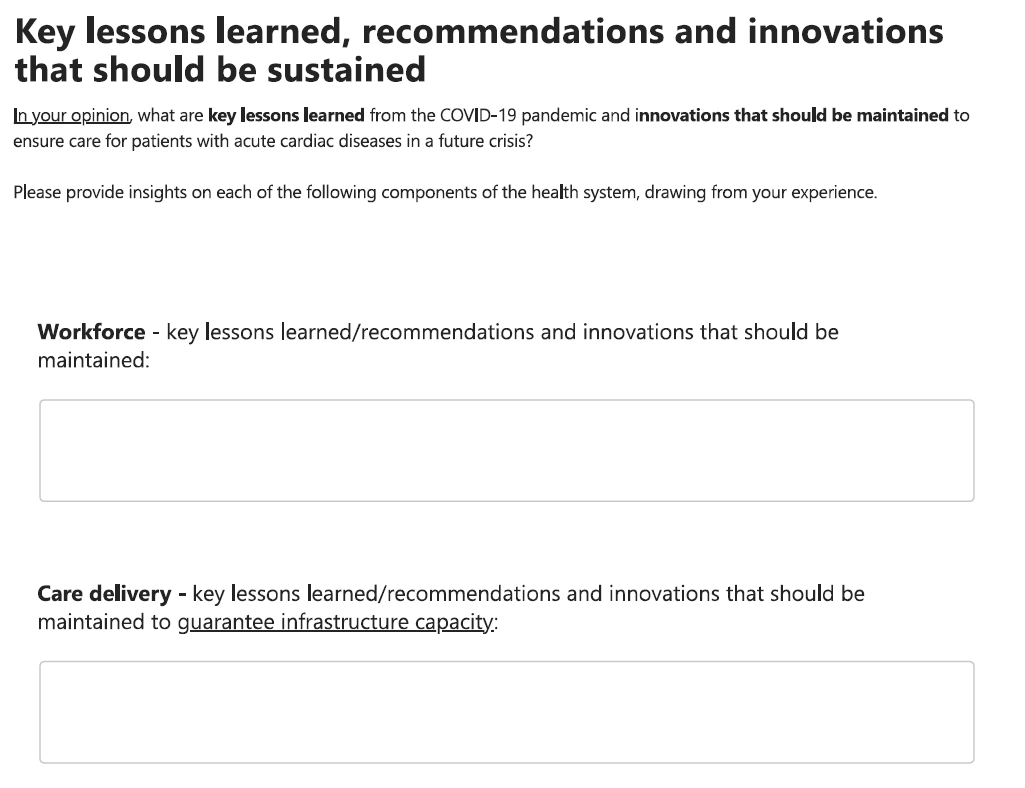

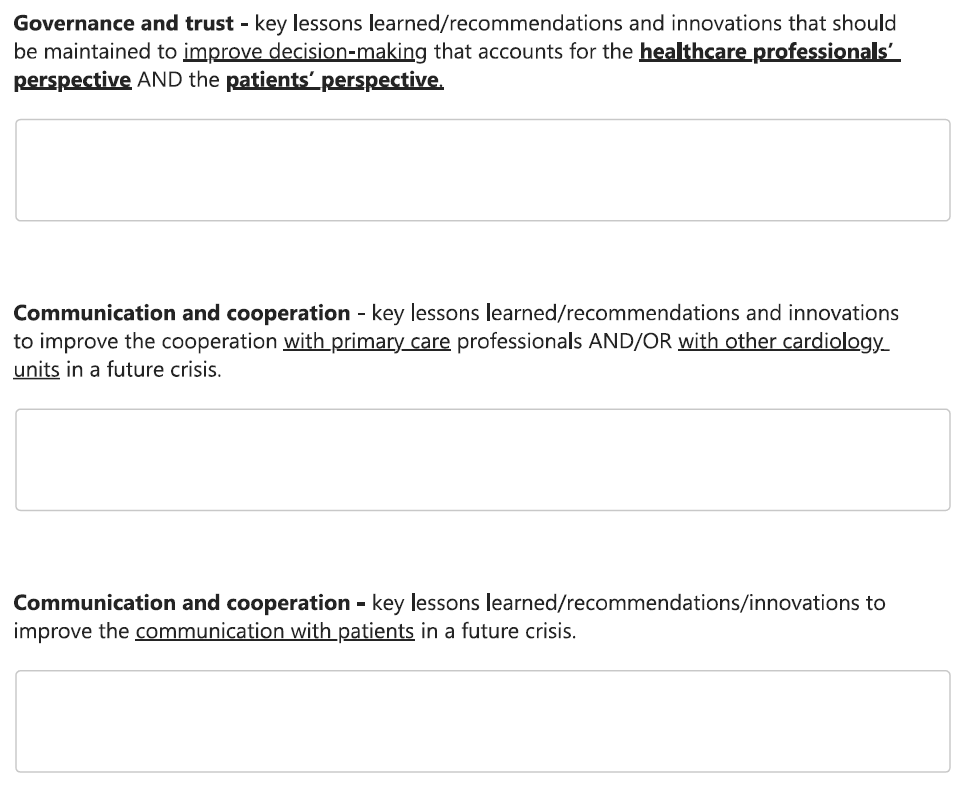

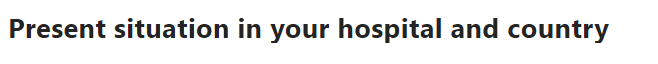

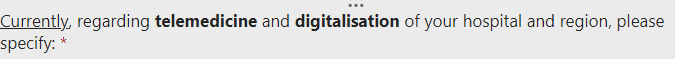

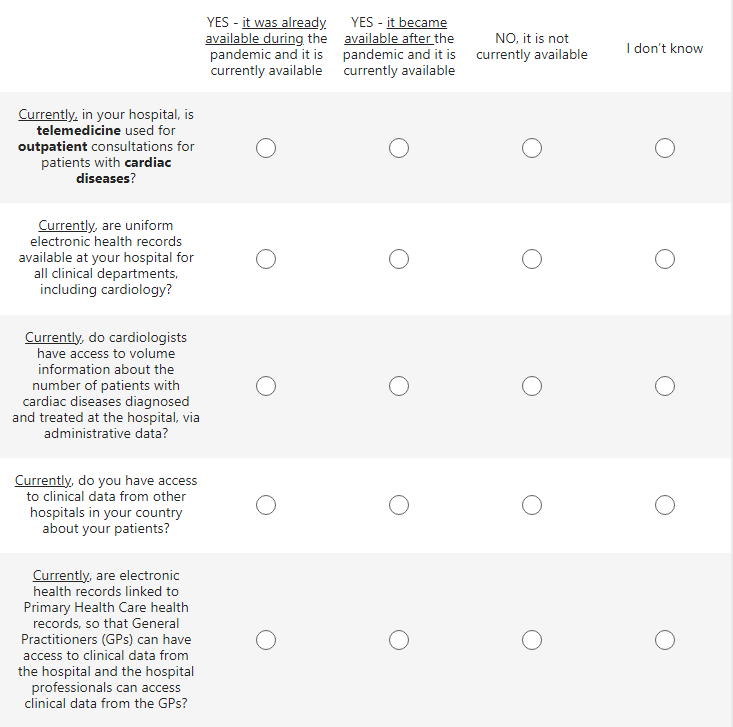

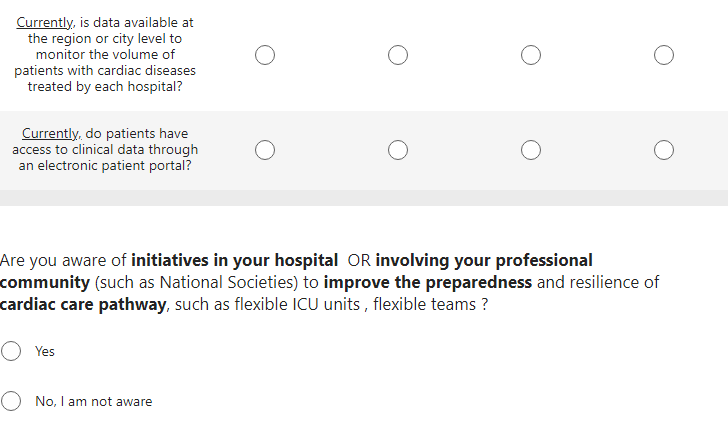

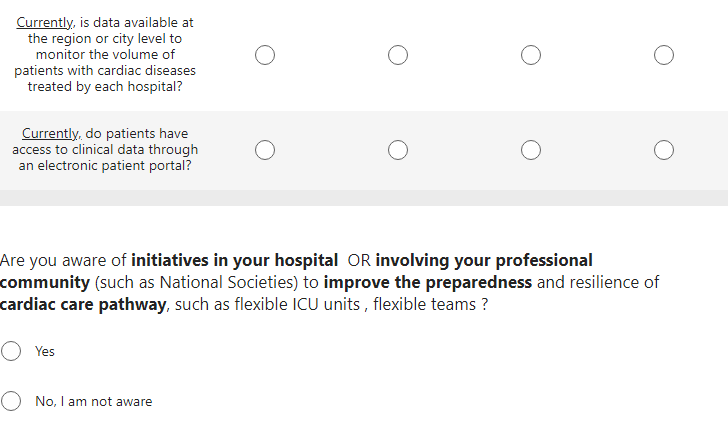

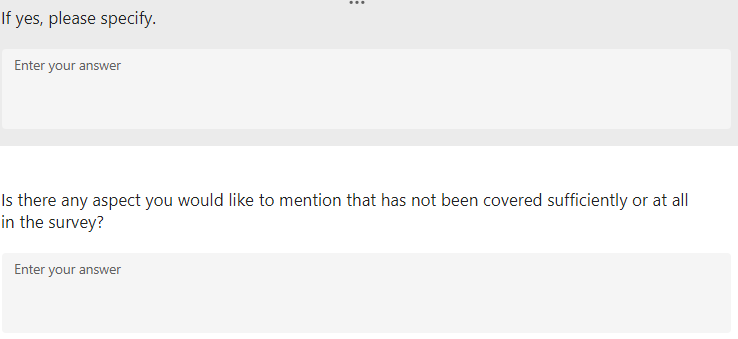

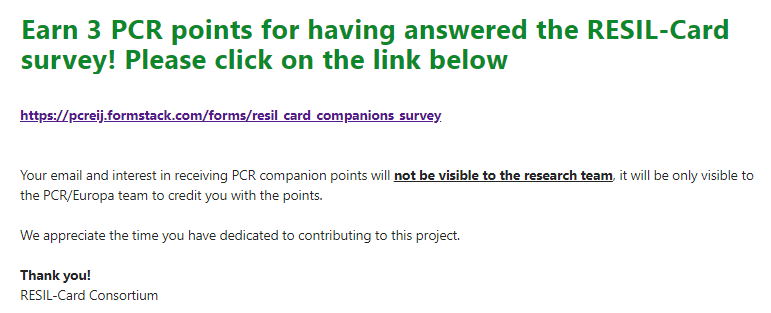
