## Supplementary material for "Development of a resilience assessment tool for cardiac care pathways in Europe: A mixed-methods study": Organisations and initiatives to raise awareness to the RESIL-Card questionnaire

### Supplementary file 4 - Organisations and initiatives to raise awareness to the RESIL-Card questionnaire (May and June 2024)

Several partner organisations were involved in the questionnaire dissemination, namely ‘Women as One’, members of the European Association of Percutaneous Cardiovascular Interventions Nurses and Allied Professionals (EAPCI NAPs) and PCR NAPs, as well as National Working Groups and Societies of Cardiology of the 27 EU MS and Ukraine.

- **Women as One:** questionnaire sent to 319 women physicians on 24-05-2024 with a reminder on the 30-05-2024;
- **Nurses and Allied Professionals (NAPs):** the survey was promoted on the LinkedIn and Facebook of NAPs groups with around 700 followers (24-05-2024)
- **National networks** in France, Spain, Portugal, Italy, UK, Greece, Netherlands, Norway and Denmark. Reminder sent on the 30-05-2024.

Initiatives to raise awareness to the RESIL-Card questionnaire (May and June 2024) are described in the table below:

| **Event, local and date** | **Initiative** |
| --- | --- |
| **EuroPCR 2024 (Paris, 2024;  14-17 May)**  4-day Course to Interventional Cardiologists in Paris, where several initiatives were developed to raise awareness to the project and to the survey  Event attended by 12,100 participants | - 2 min interview in PCR television with 2 principal investigators of the project;  - initiative to raise funds by counting steps in the congress (“count your steps for a good cause”);  - 20-min session in the PCR companions lounge by one principal investigator of the project;  - article in the daily journal of the congress (delivered in paper daily to participants) |
| **Email communication through Catalan Society of Cardiology, Spain**  15th May, 2024 | Email communication among database of cardiology professionals to encourage participation. |
| “**Best of EuroPCR” emailing list**  23^th^ May 2024 | Survey featured in the “Best of EuroPCR” emailing list, after the EuroPCR congress |
| **Reminder email to PCR mailing list**  24^th^ May 2024 | Reminder email to the EU27 and Ukraine PCR Companions |
| **RESIL-Card website update**  28^th^ May 2024 | The direct link to the survey was made available on the RESIL-Card website |
| **Email communication through National Institute for Prevention and Cardiovascular Health (NIPC), Ireland**  30^th^ May 2024 | Email communication among database of healthcare professionals to encourage participation. |
| **Email communication through Catalan Health Service, Spain 30th May, 2024** | Email communication among members of the Catalan Strategic Framework for Cardiovascular Diseases. |
| **Video with principal investigator on social media**  30^th^ May 2024 | Post on social media through various PCR channels using a short video recorded by a principal investigator explaining the importance of the survey |
| **Congress of the Catalan Society of Cardiology Barcelona University**  29^th^ - 31^st^ May 2024 | Poster in a dedicated space to the project and one of the investigators from Spain present to encourage participation |
| **Congress “Think Heart” in Italy**  30th May 2024, Rome | Presentation by one principal investigator |
| **Journal article**  “Health Management”, Volume 24 - Issue 2, 2024 | “Strengthening Cardiovascular Care Resilience for Healthier Hearts: The RESIL-Card Project (healthmanagement.org)”  Article about the aims of the project with mention to the survey |
| **PCR newsletter via email**  8 June 2024 | Reference to RESIL-Card survey in the PCR newsletter that distributed to all PCR members |
| **Social media publications**  11 and 13 June | Final reminders posted on X, LinkedIn, Instagram, and Facebook. |
| **Presentation to the Working Group on Spanish PCI meeting**  14^th^ June 2024 | Presentation by one principal investigator |
