## Supplementary material for "Development of a resilience assessment tool for cardiac care pathways in Europe: A mixed-methods study": Focus groups - Breakdown of total number of invitations and participation

### Supplementary file 5 - Focus groups - Breakdown of total number of invitations and participation in each focus group

| Country | N invited | N unavailable | N did not respond | N confirmed | N which have attended the focus group | N Absent |
| --- | --- | --- | --- | --- | --- | --- |
| Spain | 15 | 1 | 3 | 11 | 11 | 0 |
| Italy Lombardia | 11 | 0 | 0 | 11 | 10 | 1 |
| Italy Campagnia | 9 | 0 | 0 | 9 | 8 | 1 |
| NL | 9 | 2 | 3 | 4 | 4 | 0 |
| EU | 11 | 2 | 1 | 8 | 7 | 1 |
| TOTAL | **55** | **5** | **7** | **43** | **40** | **3** |
