## Supplemental Data 1 for "Development of a resilience assessment tool for cardiac care pathways in Europe: A mixed-methods study"

### Supplemental File 6 - Survey results – Study population

| **CHARACTERISTICS** | **TOTAL OF RESPONDENTS TO THE SURVEY, N=177** | |
| --- | --- | --- |
|  | **n** | **%** |
| **Role** |  |  |
| Physician | 163 | 92% |
| Nurse | 13 | 7% |
| Allied Health Professional | 1 | 1% |
| **Specialisation** |  |  |
| ***Physicians' specialisation (n=163)*** |  |  |
| Interventional Cardiologist | 124 | 70% |
| General Cardiologist | 27 | 15% |
| Cardiologist specialised in imaging | 11 | 6% |
| Cardiologist specialised in acute cardiac care | 7 | 4% |
| Cardiac surgeon | 7 | 4% |
| Cardiologist specialised in heart failure | 4 | 2% |
| Cardiologist specialised in electrophysiology | 3 | 2% |
| Ophthalmologist | 1 | 1% |
| Paediatric cardiologist | 1 | 1% |
| Cardiology Fellow | 1 | 1% |
| Cardiologist specialised in preventive cardiology | 1 | 1% |
| ***Nurses' specialisation (n=13)*** |  |  |
| Cardiac catheterisation laboratory (Cath lab) nurse | 8 | 5% |
| Cardiac care units nurse | 3 | 2% |
| Pacemaker & Implantable Cardioverter Defibrillator clinic | 1 | 1% |
| Intensive care unit (ICU) nurse | 1 | 1% |
| ***Allied professionals' specialisation (n=1)*** |  |  |
| Cardiac Catheterization Lab Technicians (Cath Lab Techs) | 1 | 1% |
| **Country** |  |  |
| Italy | 47 | 27% |
| Spain | 43 | 24% |
| Sweden | 15 | 8% |
| Greece | 12 | 7% |
| Austria | 7 | 4% |
| France | 7 | 4% |
| Belgium | 6 | 3% |
| Germany | 5 | 3% |
| Ireland | 5 | 3% |
| Netherlands | 5 | 3% |
| Portugal | 4 | 2% |
| Slovenia | 3 | 2% |
| Ukraine | 3 | 2% |
| Poland | 3 | 2% |
| Denmark | 2 | 1% |
| Luxembourg | 2 | 1% |
| Hungary | 2 | 1% |
| Bulgaria | 1 | 1% |
| Norway | 1 | 1% |
| Czechia | 1 | 1% |
| Romania | 1 | 1% |
| Croatia | 1 | 1% |
| Cyprus | 1 | 1% |
| **Sex** |  |  |
| Man | 107 | 60% |
| Woman | 69 | 39% |
| Prefer not to answer | 1 | 1% |
| **Years of professional experience** |  |  |
| ≥26 | 41 | 23% |
| 1-10 | 46 | 26% |
| 11-15 | 34 | 19% |
| 16-20 | 30 | 17% |
| 21-25 | 26 | 15% |
| **Professionals in training during the COVID-19 pandemic** |  |  |
| No | 130 | 73% |
| Yes | 47 | 27% |
| **Characteristics of the organisation** |  |  |
| Regional hospital with cath lab working 24/7 | 83 | 47% |
| Academic or non-academic referral center | 77 | 44% |
| Regional hospital without cath lab working 24/7 | 9 | 5% |
| General hospital with no specialty training for cardiologists | 8 | 5% |
| **Type of hospital** |  |  |
| Public/Government hospital | 153 | 86% |
| Private non-profit | 15 | 8% |
| Private for-profit | 9 | 5% |
