## Supplementary material for "Development of a resilience assessment tool for cardiac care pathways in Europe: A mixed-methods study": Focus groups: Detailed breakdown of informants

### Supplemental File 7 - Focus groups: Detailed breakdown of informants

| **CHARACTERISTICS** | **TOTAL OF FOCUS GROUPS INFORMANTS, N=40** | |
| --- | --- | --- |
|  | **n** | **%** |
| **Healthcare system level** |  |  |
| Micro (clinical) | 31 | 78% |
| Meso (organisational) | 4 | 10% |
| Macro (policy) | 2 | 5% |
| Cross-cutting (scientific societies, patient association) | 14 | 35% |
| **Type of stakeholder (n= 11 held two affiliations)** | | |
| Patient / patient association | 5 | 13% |
| Cardiologist | 1 | 3% |
| Head nurse (cardiology ward; cardiac care unit, cathlab) | 5 | 13% |
| Allied professional | 4 | 10% |
| Head of hospital department (cardiology, emergency department or cardiac care unit) | 10 | 25% |
| General practicioner | 4 | 10% |
| Hospital manager | 4 | 10% |
| Professional association | 15 | 38% |
| Policy-maker / governmental organisations | 3 | 8% |
| **Country** |  |  |
| Italy | 18 | 45% |
| Spain | 11 | 28% |
| Netherlands | 4 | 10% |
| Sweden | 2 | 5% |
| Romania | 1 | 3% |
| Greece | 1 | 3% |
| Belgium | 1 | 3% |
| Portugal | 1 | 3% |
| Poland | 1 | 3% |
| **Gender** |  |  |
| Male | 25 | 63% |
| Female | 15 | 38% |
