## Supplementary material for "Development of a resilience assessment tool for cardiac care pathways in Europe: A mixed-methods study": Survey results, Section1: Challenges before the COVID-19 pandemic

### Supplemental file 8 - Survey results, Section1: Challenges before the COVID-19 pandemic

66 respondents (37%) rated shortage of infrastructure capacity as a very important and extremely important challenge to care delivery before the pandemic and 64 (36%) rated nurses or physicians shortages as a relevant challenge.

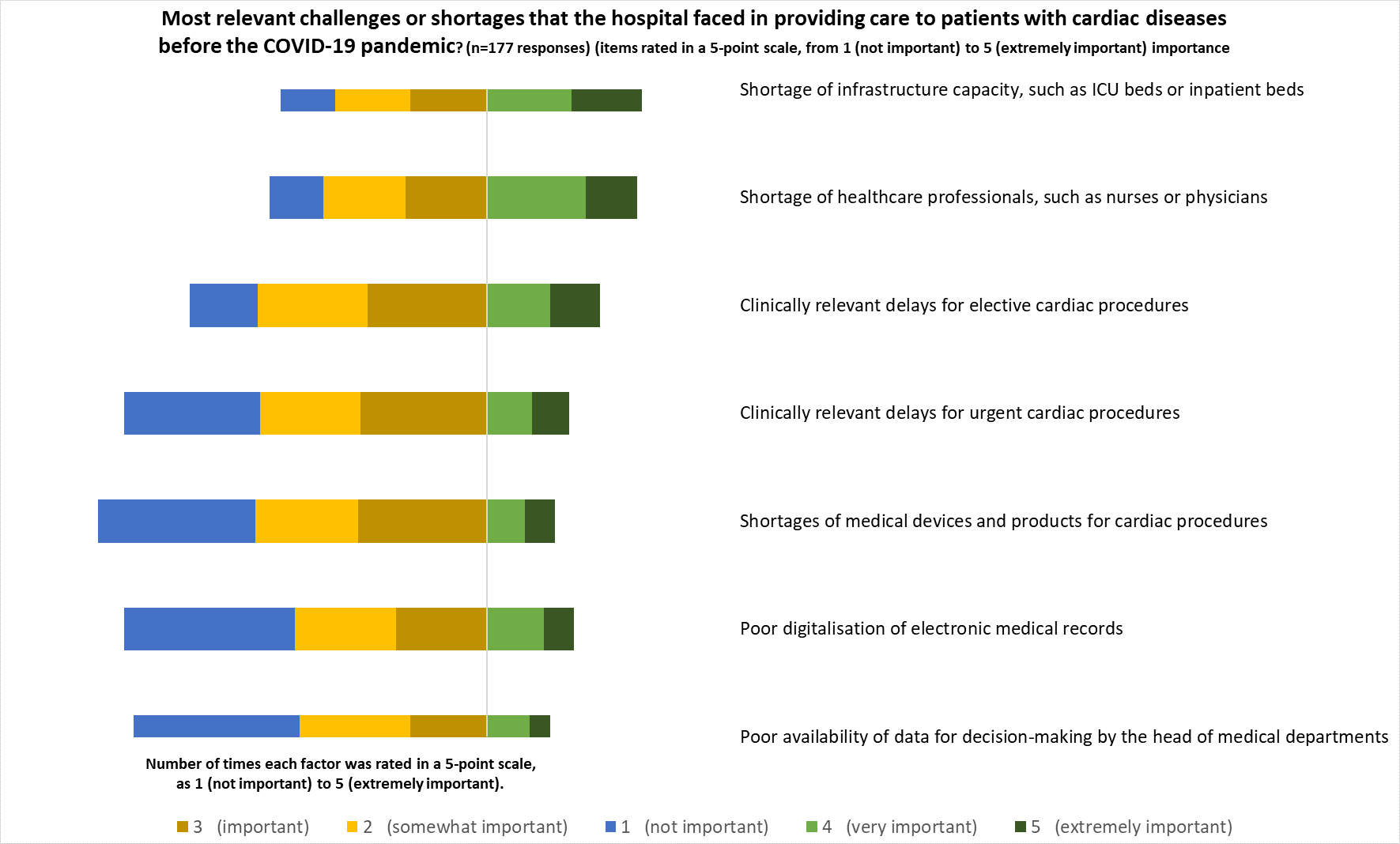
