## Supplementary material for "Development of a resilience assessment tool for cardiac care pathways in Europe: A mixed-methods study": Survey results, Section2: Nature of disruptions during the COVID-19 pandemic and the practices and innovations implemented

### Supplemental File 9 - Survey results, Section2: Nature of disruptions during the COVID-19 pandemic and the practices and innovations implemented

| Resilience dimension | Nature of disruptions during the COVID-19 pandemic | Practices and innovations |
| --- | --- | --- |
| Workforce | Impact of the shortage of healthcare professionals in care delivery classified as medium/low:  47 (x%) medium impact  45 (%) low impact | Policies implemented to address shortages of healthcare professionals (n=177):   - Reallocation health staff to key clinical areas (n=133; 75%) - Reorganisation of clinical teams (n=77; 44%) - Increase working hours (n=76; 43%) - Increase workload (n=71; 40%) - Task shifting and sharing (n=67; 38%) - Provide rapid training in key clinical areas (e.g., intensive care, primary health care) (n=51; 29%) - Recruiting new staff to support increased patient volumes in the hospital (n=38; 21%) - Reallocate health workers to other hospitals with greater needs or receiving temporary staff from hospitals (n=33;19%) - Increase digitalization of clinical data (n=16; 9%) - Recruiting non-health workers, such as post-office-workers, firefighters, army (n=10; 6%) |
| Care delivery | - The most frequent shortages in infrastructure capacity reported were related to ICU beds (n=152, 73.4%). - Overall, Emergency Medical Services ambulances were well equipped during the COVID-19 pandemic - Delays in accessing diagnostic procedures were frequently reported (66%; n=116)   The most frequent reasons for delays in accessing diagnostic procedures were (n=177; multiple responses allowed):  - Decision, at hospital level, to stop providing specific procedures (n=69; 39%);  - Mobilisation of human and/or material resources (e.g. personal protective equipment) to other departments (n=63; 36%);  - Lack of human resources for individual reasons, such as quarantine for COVID-19 infection (n=52; 29%).   - The treatment procedures where waiting times were most frequently reported, from a clinical perspective, during the COVID-19 pandemic, were: elective diagnostic catheterization (n=120; 68%) and elective invasive coronary angiography/PCI for chronic coronary syndromes (n=112;63%), followed by TAVI (n=105; 59%), ablation for AF (n=104; 59%), and surgical aortic valve replacement (n=103;58%). - The most frequent reasons to restrict access to urgent treatment were: - infrastructure capacity shortage (cath lab, operation room, ICU...): 38% rated it as very important and extremely important; - shortage of healthcare professionals because of quarantine or COVID-19 diagnosis: 35% rated it as very important and extremely important and - health protection of professionals (to minimize exposure of providers to the COVID-19 infection): 28% rated it as very important and extremely important. | The actions taken to address shortages in infrastructure capacity at the hospital level (n=85 responses):   1. New Infrastructures 2. Temporary Infrastructures or repurposing of previous infrastructures 3. National and International Collaboration   New care delivery models for patients with cardiac diseases since the start of the pandemic were reported by 66 of the respondents (37%), while 96 (54%) reported that there were any innovation implemented. |
| Communication | - Changes in referral policies were frequently reported (n=95/177; 54%). The most frequent change in referral policy was ‘referral criteria between primary health care and hospital care and end-of-life care’ (n=61/109, %) | - New communication channels to inform patients were implemented in almost half of the cases (n=76; 43%).   The most common channels used were the phone (n=50 out of 154 responses; 32%), followed by the email (n=36 out of 154 responses; 23%) and information via social media (n=24; 16%)   - Public awareness campaigns to citizens were frequent (n=101; 57%). |
| Governance | The professionals most frequently involved and responsible for decision-making were hospital managers and the heart team, with the percentages depending on the decisions in analysis:  - the professionals most frequently involved in decisions related treatment options for specific patients and prioritisation of patients for treatment were the heart team (reported by 61% and 48% of respondents, respectively)  - the professionals most frequently involved and most frequently responsible for decisions related to task substitution were hospital managers (n=98; 55%; n=68; 38%)  - the professionals most frequently responsible for decisions related to changes in care delivery were Head of cardiology department (n=80; 45%) | How professionals were involved in the decisions regarding prioritisation of patients (n=177): most respondents reported consensus decision-making through meetings/doctors as party of decision committees (n=98/177; %), while 63/177 (%) reported individual consultation with some professionals.  A diversity of cardiology guidelines provided guidance during the COVID-19 pandemic, including international guidelines, national, regional and some hospital/local guidelines. |
| Data | Data was available to monitor the volume of patients with cardiac diseases treated by each hospital in almost half of the responses (n=81; 45.8%) |  |
| Devices | Shortages of essential medical devices reported in almost half of responses (86; 49%), and those shortages were mostly related to mechanical ventilators (n=77 out of 163 responses; 47% ). |  |
