## Supplementary figures and images for "Development of a resilience assessment tool for cardiac care pathways in Europe: A mixed-methods study"

### Survey results, Section3: Lessons learned after the COVID-19 pandemic

#### Supplemental file 10 - Survey results, Section3: Lessons learned after the COVID-19 pandemic

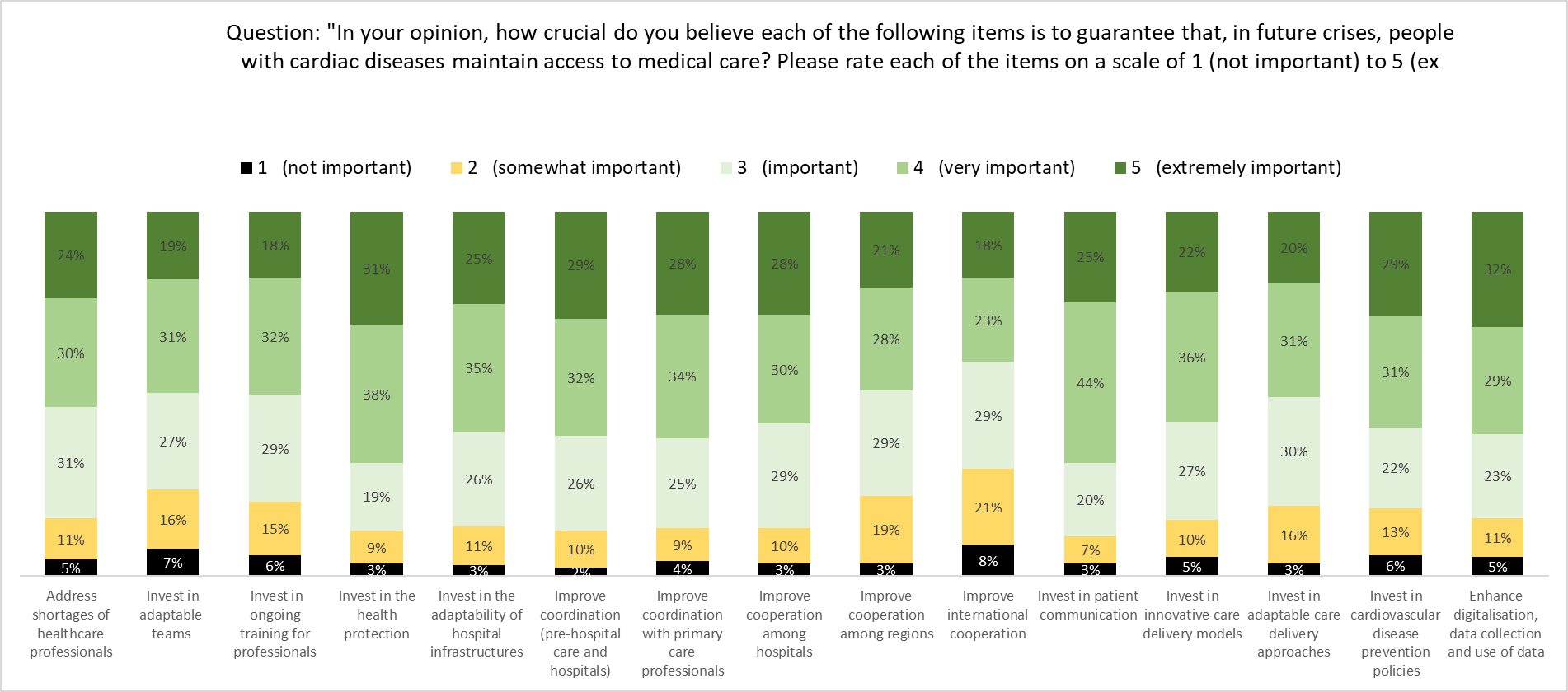
