## Supplementary material for "Development of a resilience assessment tool for cardiac care pathways in Europe: A mixed-methods study": Themes identified to develop the resilience tool, per resilience dimension

### Supplementary file 11 - Themes identified to develop the resilience tool, per resilience dimension

Survey themes were complemented and refined with the focus groups discussions, per resilience dimension, to develop the resilience tool

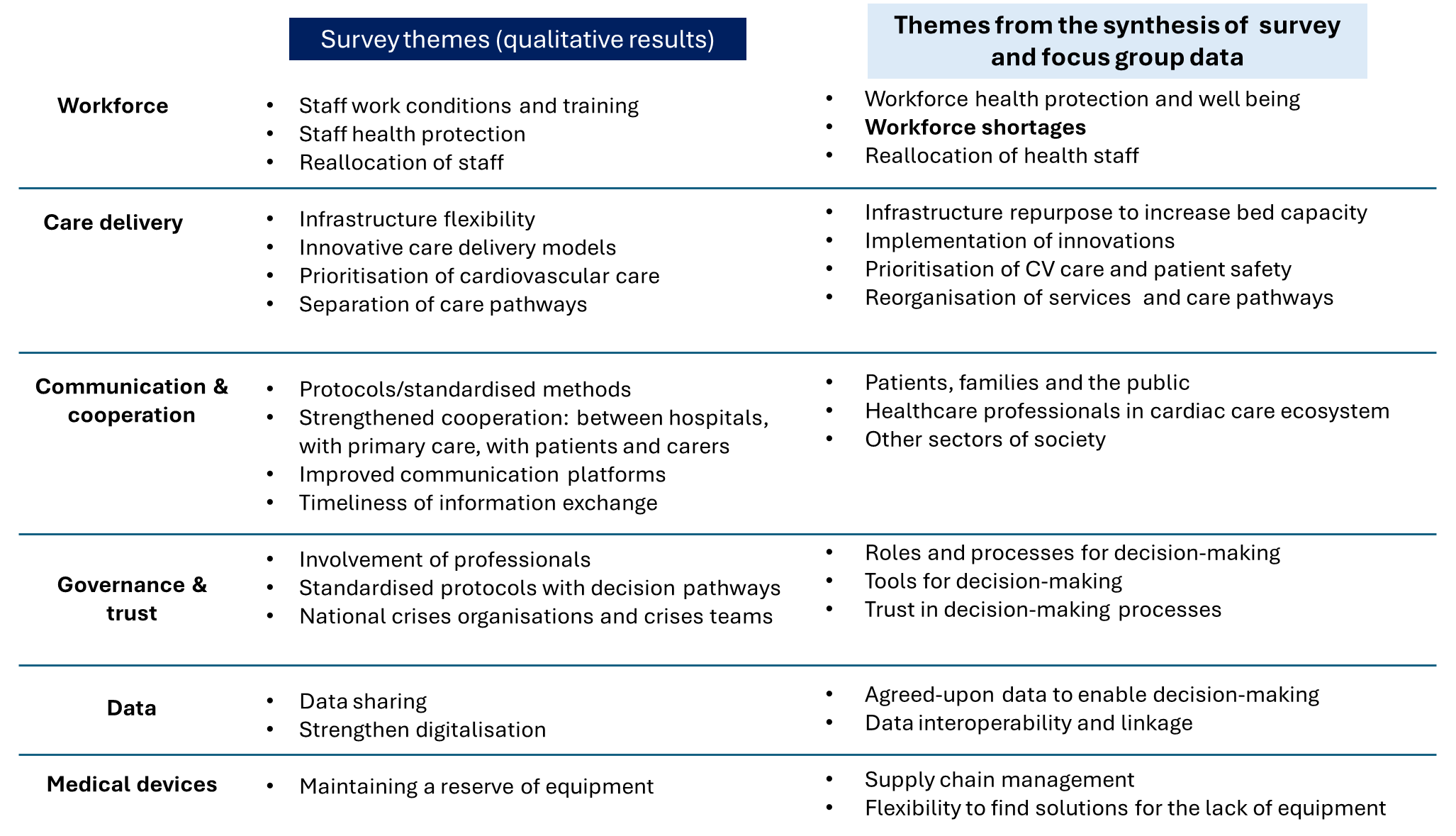
